## Supplementary Information for "Digital proximity tracing on empirical contact networks for pandemic control"

[1] *Fondazione Bruno Kessler, Trento, Italy*
[2] *University of Trento, Trento, Italy*
[3] *University of Padua, Padua, Italy*
[4] *Aix Marseille Univ, Université de Toulon, CNRS, CPT, Turing Center for Living Systems, Marseille, France*
[5] *Tokyo Tech World Research Hub Initiative (WRHI), Tokyo Institute of Technology, Tokyo, Japan*
[6] *University of Turin, Turin, Italy*
[7] *ISI Foundation, Turin, Italy*
[8] *Technical University of Denmark, Copenhagen, Denmark*
[9] *École Polytechnique Fédérale de Lausanne (EPFL), Lausanne, Switzerland*

[†] *These authors contributed equally to this work.*

#### Supplementary Note 1 Characteristic parameters of the disease

In this section we provide details on the various parameters that represent the epidemic spread in both the continuous model and the network model. Moreover, we demonstrate that the model is robust with respect to the choice of the infectiousness probability as a function of the time since infection.

##### Supplementary Note 1.1 Infectiousness parameters in the continuous model

The choice of the infectiousness function and the epidemic parameters that describe the COVID-19 spreading in the continuous model follow the work of Ferretti et al. [1], with some modifications that we describe here and summarize in Supplementary Table 2.

The infectiousness $\omega(\tau)$ is a function of the days since infection, proposed by Ferretti et al. [1]. It takes into account four different contributions: asymptomatic, pre-symptomatic and symptomatic infectiousness, plus environmental transmission representing the indirect contagion occurring for instance via contaminated surfaces. The symptomatic infectiousness has been obtained by Ferretti et al. by making use of generation time data. The pre-symptomatic infectiousness is assumed to be equal to the symptomatic one, while the asymptomatic individuals are considered to have only 10% of the infection potential, according to the recent literature [2, 3]. An alternative shape of the curve $\omega(\tau)$ is discussed in Supplementary Note 1.3. The infectiousness is a probability distribution and as such it is normalized to one. It appears in the model equation (1) in the main text multiplied by $R_0$, that we consider equal to 3 when no measure is implemented. All the analyses are however performed using reduced values, $R_0 = 1.2, 1.5, 2.0$, which take into account the combined effect of all the alternative measures (masks, physical distancing, etc.) in a range suggested by recent literature [4, 5, 6, 7].

For the cumulative distribution $s(\tau)$ of onset times (i.e. time between infection and appearance of symptoms), we adopt the assumptions of Ferretti et al. [1] with two modifications. This function actually gives the fraction of the infected population that becomes known as infected by the health authorities, and does not distinguish between symptomatic individuals and asymptomatics identified by randomized testing. This is the same assumption as in Ferretti et al. [1], and it is motivated by the fact that the tracing and quarantining policy is activated independently of the source of knowledge of the infected status. The first modification to the onset time is that we rescale the function $s$ so that its cumulative probability $s(\tau)$ reaches $p = 0.8$ at large times instead of 1. This models our assumption that even at infinite time only 80% of the infected population is detected, instead of 100%. This describes a situation in which 60% of infected are symptomatic, and additionally 50% of asymptomatics are identified by randomized testing, or equivalently to a situation with 80% symptomatics and no randomized testing. The second modification is that we shift the symptom onset forward in time by 2 days, modelling a delay in the functioning of the testing and reporting policy. Different assumptions on this delay are discussed in Supplementary Note 3.2.

#### Supplementary Note 1.2 Parameter tuning to validate the infection probabilities

As mentioned in the main text in Section , the CNS data set provides us with the opportunity to explore the dependence of the infectiousness from duration and proximity, a question to which the literature is not yet able to express a specific answer. We rely on some simplifying assumptions by supposing that in occasion of a contact between an infected and a susceptible person the contagion probability depends only on their proximity, on the duration of the contact and on the time since the infectious individual has been infected. We moreover assume that those probabilities are independent from each other and require that, if simulated on the CNS data set without any restriction, the resulting reproductive number is equal to $R_0 = 3$, in agreement with recent literature on the COVID-19. Given a choice of the infectiousness parameters, the corresponding value of

| Name | Inputs | Definition | Description |
|---|---|---|---|
| $\omega(\tau)$ | time $\tau$ (days) | Weibull distribution with shape = 2.826 and scale = 5.665. | Probability for an infected individual to transmit the disease at time $\tau$. |
| $R_0$ | | 1.2, 1.5, 2 | Reproductive number. |
| onset_time($\tau$) | time $\tau$ (days) | Lognormal distribution with $\mu = 1.54$, $\sigma = 0.47$, shifted by the delay of 2 days, and scaled in $[0, 0.8]$. | Probability for an infected individual to be detected exactly at time $\tau$. |
| $s(\tau)$ | time $\tau$ (days) | Cumulative distribution of onset_time($\tau$). | Probability for an infected individual to be detected within time $\tau$. |

Supplementary Table 2: Characteristic parameters of the disease that are used in the continuous model.

$R_0$ is estimated by computing an empirical value $R_0^{data}$. This is obtained by numerically simulating the epidemic spreading, assuming one random individual initially infected, and counting the number of secondary infections caused by this patient zero [8]. The average of this value over multiple independent runs is the estimated value $R_0^{data}$.

The infectiousness function is thus defined as:

$$\omega_{\text{data}}(\tau, e, s_s) = r_{R_0} \cdot p_{R_0} \cdot \omega(\tau) \cdot \omega_{\text{exposure}}(e) \cdot \omega_{\text{dist}}(s_s) \tag{1}$$

where $\omega(\tau)$ is the probability for an infected individual to transmit the disease at time $\tau$ after its own infection, $\omega_{\text{exposure}}(e)$ is the probability to transmit the disease given the duration $e$ of a contact, and $\omega_{\text{dist}}(s_s)$ is the probability as a function of the signal strength $s_s$ of the contact. The constant $r_{R_0}$ is a reduction factor that can be tuned to obtain the desired value of $R_0$, and $p_{R_0}$ is a scaling factor. Using two distinct scaling factors allows us to decouple the estimate of the parameters to obtain the target value of $R_0 = 3$, and the computation of the reduction factor needed to obtain a smaller value.

Considering everything fixed except for $\omega_{\text{exposure}}(e)$ and $\omega_{\text{dist}}(s_s)$ we can play with the free parameters of these functions so as to explore different scenarios while keeping a balance between time and space dependencies corresponding to an $R_0$ around 3 (with $r_{R_0} = 1$).

The shape of $\omega_{\text{exposure}}$ has been inspired by the literature [9, 8, 10]:

$$\omega_{\text{exposure}}(e) = (1 - \beta_0)^{e/dt}, \tag{2}$$

where $dt$ is a time step and $\beta_0$ a free parameter. The value of $\beta_0$ can be set by requiring that a specific probability $\sigma$ for an infected individual to transmit the disease is reached for a given contact duration $e_\sigma$:

$$\omega_{\text{exposure}}(e_\sigma) = \sigma. \tag{3}$$

The parameter $\beta_0$ can thus be expressed as a function of $e_\sigma$ and $\sigma$ as:

$$\beta_0\left(e_\sigma, \sigma\right) = 1 - \left(1 - \sigma\right)^{dt/e_\sigma}. \tag{4}$$

Supplementary Table 3 reports some examples. For instance, to obtain a 90% probability of infection for contacts of 1 hour, the parameter $\beta_0$ needs to be set equal to 0.038.

| | $e_\sigma$ $[hours]$ | $\sigma$ | $\beta_0$ |
|---|---|---|---|
| ● | 1.0 | 0.9 | 0.038 |
| ● | 2.0 | 0.9 | **0.019** |
| ● | 4.0 | 0.9 | 0.010 |

Supplementary Table 3: Numerical values for $\beta_0$ for three different sets of physical scenarios $(e_\sigma, \sigma)$. The value of $\beta_0$ highlighted in bold is the one chosen for the simulations reported in all the other sections.

The term $\omega_{\text{dist}}(s_s)$ instead depends on the Bluetooth signal strength (RSSI), expressed in dBm, which is considered as a proxy for the distance between individuals. We thus define the function $\tilde{\omega}(x) = \omega_{\text{dist}}(s_s(x))$, where $x$ indicates distances in meters. We emphasize here again that the relationship between RSSI and distance is far from trivial [11, 12], so in the main text we will rely on signal strength as a proxy for distance.
To our knowledge, the literature on COVID-19 has not yet produced some evidence regarding the probability of contagion as a function of the distance between an infected individual and a susceptible one. We make the realistic assumption that infectiousness is large when the individuals are in close proximity and that it decreases with distance. In particular we hypothesize that it follows a sigmoid function:

$$\tilde{\omega}_{\text{dist}}(x) = \frac{s}{\log\left(1 + \mathrm{e}^{b}\right)} \left(1 - \frac{1}{1 + \mathrm{e}^{b - sx}}\right), \tag{5}$$

where $s$ and $b$ are free parameters. As we have two parameters, we need to specify two physical conditions to find their values. We then require that the probability for an infected individual to transmit the disease to a contact within a distance $x_i$ $(i = 1, 2)$ should be $w_i$ $(i = 1, 2)$:

$$\begin{cases} \int_0^{x_1} \tilde{\omega}_{\text{dist}}(x) dx = w_1 \\ \int_0^{x_2} \tilde{\omega}_{\text{dist}}(x) dx = w_2 \ . \end{cases} \tag{6}$$

Computing explicitly the integrals using Eq. (5), we obtain

$$\begin{cases} 1 - \frac{\log\left(1 + \mathrm{e}^{b - sx_1}\right)}{\log\left(1 + \mathrm{e}^{b}\right)} = w_1 \\ 1 - \frac{\log\left(1 + \mathrm{e}^{b - sx_2}\right)}{\log\left(1 + \mathrm{e}^{b}\right)} = w_2 \end{cases} \tag{7}$$

which is a transcendental system, that can be numerically solved once we have set the two couples $(x_i, w_i)_{i=1,2}$. Some examples are reported in Supplementary Table 4.

| | $x_1$ $[m]$ | $w_1$ | $x_2$ $[m]$ | $w_2$ | $s$ $[m^{-1}]$ | $b$ |
|---|---|---|---|---|---|---|
| ● (blue) | 1.7 | 0.5 | 6.0 | 0.99 | 1.16 | 3.65 |
| ● (orange) | 2.5 | 0.5 | 7.0 | 0.99 | **1.34** | **6.67** |
| ● (pink) | 4.0 | 0.5 | 10.0 | 0.99 | 1.16 | 9.31 |

Supplementary Table 4: Numerical solutions $(s, b)$ for the system (7) for three different sets of physical requests $(x_i, w_i)_{i=1,2}$. The values of $s$ and $b$ highlighted in bold are the ones chosen for the simulations reported in all the other sections.

The three curves that we obtain using the values in Supplementary Table 3 and Supplementary Table 4 are shown in Supplementary Fig. 1.

While the reproductive number of COVID-19 is estimated to be around 3 [13], there is small evidence for the dependence on proximity and duration. Therefore, we combine the two functions $\omega_{\text{exposure}}(e)$ and $\tilde{\omega}_{\text{dist}}(x)$ and choose the parameters $\beta_0$, $b$ and $s$ to obtain $R_0 = 3$ in each combination. In particular, given a possible choice for $(\beta_0, b, s)$, we run a set of 800 simulations on the CNS data set without any restrictive policy, i.e. with $\varepsilon_I = 0$ and one initial infected. We then count the number of secondary infections caused by this first individual and average this number on all the 800 simulations to obtain an estimate of $R_0$. The constraint $R_0 = 3$ requires to find a balance between $\omega_{\text{exposure}}$ and $\tilde{\omega}_{\text{dist}}$ and combine the parameters accordingly. If for instance we suppose that infectiousness decreases slowly even at long distances (like in the last row of Supplementary Table 4) we should set $\beta_0$ such that the infectiousness of contacts has a slow increase with duration (like in the last row of Supplementary Table 3), in order not to have a huge $R_0$, and we obtain the pink curves in Supplementary Fig. 1. Vice-versa, if $\tilde{\omega}_{\text{dist}}$ is adjusted such that only close contacts are contagious, we should give more importance to duration and suppose that also short durations are at risk (e.g. blue curves in Supplementary Fig. 1).

In the numerical simulations discussed in the main text, we use the intermediate curves in Figure 1 (in orange) as infectiousness functions. We report in Supplementary Fig. 2 some results obtained by using in the simulations the two other sets of curves. The left and central panels represent the growth or decrease of the epidemic with the different policies assuming respectively the pink curves (thus assuming that contagion can take place even at long distance but only for long contact duration) and the blue ones (assuming contagion even for short durations but only at close proximity). We observe that for what concerns the controllability of the epidemics the two choices of proximity-duration dependence of infectiousness do not bring significantly different results. Nevertheless, the right panel in Supplementary Fig. 2 shows effectiveness and cost of each policy for the three proposed curves of infectiousness, and we notice that circles and diamonds have a similar trend (respectively corresponding to orange and blue curves in Supplementary Fig. 1), the choice

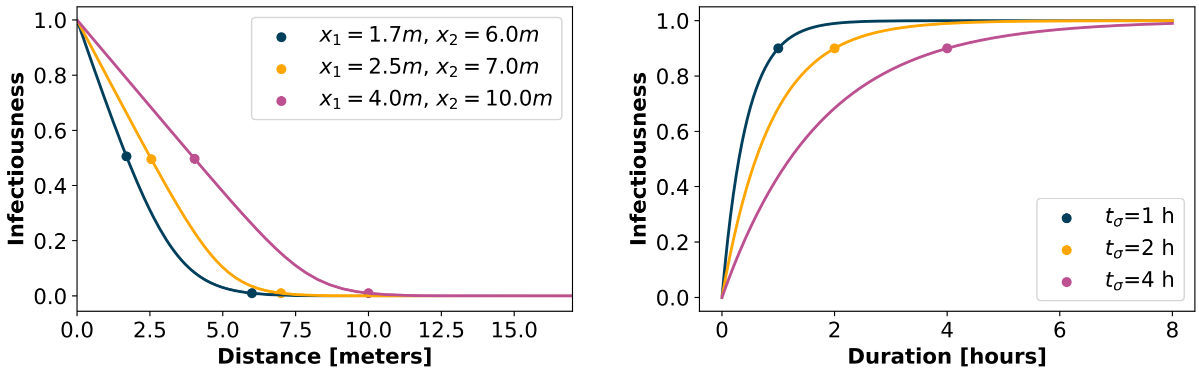


Supplementary Figure 1: Infectiousness as a function of distance (left panel) or duration (right panel) of the contact, for three different parameters configurations. By combining the two curves corresponding to each color we obtain $R_0 = 3$ in each case. The blue configuration implies an infectiousness increasing rapidly with duration but decreasing fast with distance. On the contrary, the pink curves correspond to an infectiousness that increases slowly with contact duration but has a broader spatial range. All the simulation results in the manuscript are obtained assuming the infectiousness to be ruled by the intermediate orange configuration.

of the pink curve (triangular symbols) would lead to a more optimized balance between cost and effectiveness, with lower numbers of both false negatives and total quarantined for each policy. This strengthens the idea that a better knowledge of infectiousness as a function of duration and proximity of contacts would be fundamental to devise appropriate policies to fight the pandemic.

It is worth mentioning the two constant factors $p_{R_0}$ and $r_{R_0}$ that appear in Eq. (1). The first one is just a scaling factor, that we fix to the same constant value in all settings. The second one instead plays a pivotal role. Indeed, the procedure described above for parameters' setting is aimed to reconstruct a scenario without restrictions, where the epidemic of COVID-19 is free to spread with $R_0 = 3$. In this work, we analyze the effect of isolation and tracing in a context where other protective measures contribute to mitigate the spreading. These general precautions are described in our model as an overall reduction of $R_0$, obtained by using the reduction factor $r_{R_0} \in [0, 1]$, with values reported in Supplementary Table 5. The chosen reduced values of $R_0$ take into account the combined effect of all the alternative measures in a range suggested by recent literature [4, 5, 6, 7].

Let us notice that the two functions $\omega_{\text{exposure}}$ and $\omega_{\text{dist}}$ are in principle defined as two independent functions reflecting respectively the dependency from duration and proximity. We however chose to set their free parameters simultaneously combining these two effects so as to explore how their mutual contributions change in shaping the contagions, while keeping $p_{R_0}$ fixed.

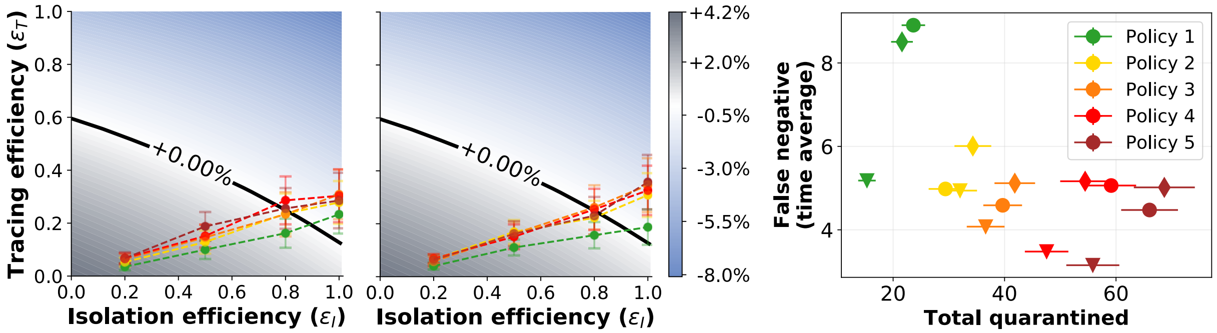


Supplementary Figure 2: Left and central panels: Growth or decrease rate of the number of newly infected individuals for each policy, assuming respectively that the dependence of infectiousness from duration and proximity follows the pink curves and the blues curves of Supplementary Fig. 1. The reducing factor $r_{R_0}$ is set to have $R_0 = 1.5$ and we assume 40% app adoption. All the points have been obtained as mean values over $n = 200$ simulations and the error bars represent the standard error. Right panel: corresponding average values of false negatives vs total quarantines for the different policies assuming for infectiousness the curves in pink (triangles), in orange (circles), and in blue (diamonds) of Supplementary Fig. 1.

| $R_0$ | 3.0 | 2.0 | 1.5 | 1.2 |
|---|---|---|---|---|
| $r_{R_0}$ | 1.0 | 0.53 | 0.39 | 0.26 |

Supplementary Table 5: In the first row the desired values of $R_0$ are reported, while the second row shows the corresponding values of the reduction factor $r_{R_0}$ needed to obtain them, with a scaling factor $p_{R_0} = 60$.

#### Supplementary Note 1.3 Robustness of the model with respect to the definition of the infectiousness probability

We consider here another infectiousness curve that has been derived in the recent literature by He et al. [14] . We follow here the author-correction version [15], that followed a critic and correction suggestion [16] on the first version.

We show that, although this curve is different from the curve $\omega$ that we use in this paper, the predictions of the model do not change significantly, showing their robustness with respect to changes in the infectiousness curve.

In the cited works the infectiousness is defined by means of two probability density functions (PDFs): The incubation time $g(t)$ (probability of symptom onset as a function of the time $t$ since infection), and the infectiousness probability $f(t)$, which is a function of the time $t$ elapsed since the symptom onset ($t$ can take negative values because of pre-symptomatic infectiousness). In more details, the function $g$ is in turn taken from Li et al. [17], and it is a lognormal distribution with mean 1.434065 and std 0.6612. The function $f$ is instead estimated by He et al. [15]: it is assumed to be a gamma distribution, and via a max-likelihood approach it is estimated to have shape 20.516508 and scale 1.592124, and to be shifted by an offset 12.272481. A numerical PDF of the two distributions, computed over $10^5$ samples, and the analytical expression of the two PDFs are shown in Supplementary Fig. 3a.

From these $g, f$, we can reconstruct a PDF $\omega_{\mathrm{He}}(\tau)$ to be used in our model. This can be done simply by sampling two values from $g$ and $f$ and adding them (the total time from infection to secondary infection is simply split into two intervals separated by the time of symptoms onset). A numerical PDF of this distribution $\omega_{\mathrm{He}}$, computed over the same $10^5$ samples, is in Supplementary Fig. 3b. This function $\omega_{\mathrm{He}}$ may also be obtained analytically by convolution as

$$\omega_{\mathrm{He}}(\tau) = \int_{-\infty}^{\infty} f(\tau - t) g(t) dt,$$

using the analytically known $f$ and $g$. The discretized convolution is also shown in Supplementary Fig. 3b, and it coincides indeed with the numerical values of $\omega_{\mathrm{He}}$.

Observe that this distribution assigns a positive probability (6.01%, see below) also to infectiousness at negative times (i.e. an individual may infect another one before being itself infected). We assume that this is due to the fact that the two distributions $f$ and $g$ are estimated from two different populations [15], and thus statistical errors may be present. For our aims this is not a limitation, as it just mean that the (cumulative) probability of infection at zero is strictly positive.

Supplementary Fig. 3b shows also the PDF $\omega$ that we used in the paper. Both distributions peak roughly at the same time ($\omega$ at 5 days, while $\omega_{\mathrm{He}}$ at 4 days). On the other hand, $\omega_{\mathrm{He}}$ has a wider support and a larger right tail, meaning that it models a non negligible probability of secondary infection also several days after the infection of the spreader.

To have an analytical expression of $\omega_{\mathrm{He}}$ we try to fit shifted lognormal, gamma, and Weibull distributions to $\omega_{\mathrm{He}}$ by least-squares minimization over the PDF obtained by convolution. The best results are obtained with a gamma distribution with density $h(\tau) = \frac{p_2^{p_1}}{\Gamma(p_1)} \tau^{p_1 - 1} e^{-p_2 \tau}$ with parameters $p_1 = 5.73$, $p_2 = 0.55$, and shifted by 4.67, which is plotted in Supplementary Fig. 3c. This allows also to derive an explicit cumulative density function $\mathrm{CDF}_{\mathrm{He}}$ of $\omega_{\mathrm{He}}$, which gives an estimate of $\mathrm{CDF}_{\mathrm{He}}(0) = 0.0601$ (the fraction of negative-time infections).

We can now use this modified infectiousness $\omega_{\mathrm{He}}$ in our model and compare the results with the ones of Fig. 5 of the main text. First, we estimate again the reduction parameter defining $\omega_{\mathrm{data}}$ (see Section  of the main text), and we get $r_{R_0} = 0.35$.

Using this functional form of $\omega_{\mathrm{He}}$ in the model, we obtain the results of Supplementary Fig. 4 (see central panel in Fig. 5 of the main text for the corresponding results with $\omega$). It is clear that the difference is quite limited since only Policy 1 and Policy 2 for

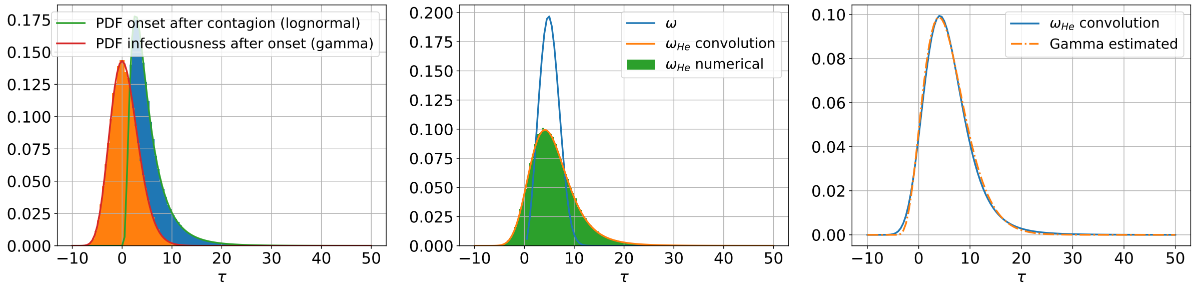


(a) (b) (c)

Supplementary Figure 3: Visualization and estimation of the infectiousness probability density function (PDF) $\omega_{\mathrm{He}}$. PDFs $f$ and $g$ (Supplementary Fig. 3a); estimated PDF $\omega_{\mathrm{He}}$, and PDF $\omega_{\mathrm{He}}$ (Supplementary Fig. 3b); fit of $\omega_{\mathrm{He}}$ with a gamma distribution (Supplementary Fig. 3c).

$\varepsilon_I = 0.8$ move from being ineffective (Fig. 5, main text) to being effective. We can thus conclude that no significant change in our conclusions would be introduced by adopting this alternative infectiousness function in place of the current one. In particular, the predictions using $\omega$ appear to be less optimistic in the prediction of the policies' effectiveness, since they estimate that not all policies are successful for $\varepsilon_I = 0.8$.

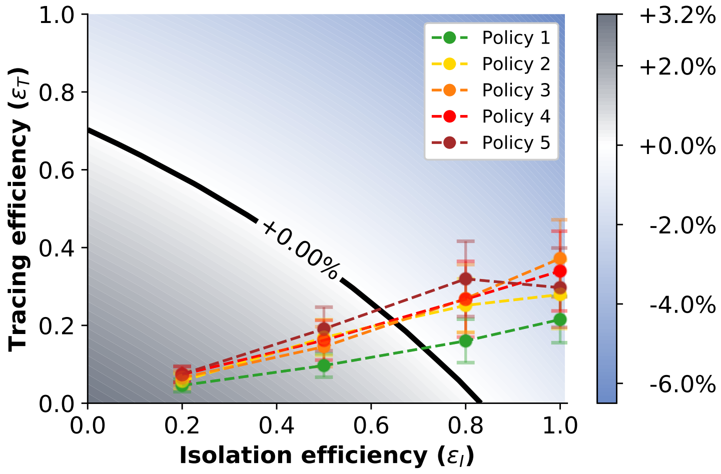


Supplementary Figure 4: **Tracing policy efficiency for alternative infectiousness.** Growth or decrease rate of the number of newly infected individuals using the modified infectiousness curve $\omega_{\mathrm{He}}$. The points correspond to the parameter pairs such that $\varepsilon_I$ is an input and $\varepsilon_T$ an output of the simulations on real contact data, for the policies of Fig. 3. Here $R_0 = 1.5$ with 40% app adoption. All the points have been obtained as mean values over $n = 200$ simulations and the error bars represent the standard error.

#### Supplementary Note 1.4 Contact patterns in the CNS data set

To further guarantee the reproducibility of the results of this paper, we provide additional details on the CNS data set.

As mentioned before, the CNS data set [18] contains one month of data that is used here as it is. Thus, for any detail we refer to the cited paper, and we only visualize in Supplementary Fig. 5a the temporal distribution of the total number of contacts contained in the data set. It is immediate to observe that the number of contacts has a periodical behavior that reflects the day/night periods and the days of the week. Moreover, a certain uniformity is present between different weeks.

For the simulations discussed in SI Supplementary Note 3.6 we need to use a longer time period, that is extracted from data that are not publicly shared in the CNS data set [18]. We extract the period from the 1st of September to the 30th of November 2013, and remove the week between 7th and 13th of October, since it corresponds to a holiday week with very few contacts. In this way, the whole timespan used for the simulations has an amount of contacts that remains on average homogeneous in time. Supplementary Fig. 5b shows the distribution of contacts in this case.

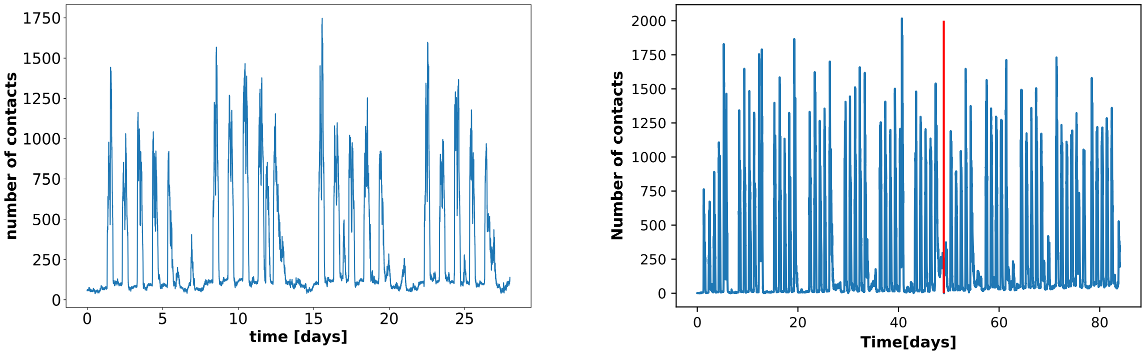


(a) (b)

Supplementary Figure 5: **Temporal distribution of the total number of contacts in the CNS data set.** The figures show the total number of contacts in the CNS data set (Supplementary Figure 5a), and in the extended version (Supplementary Figure 5b) as a function of time. The vertical red line represents the cut of the holiday week. The aggregation is computed with a temporal gap of 300 seconds.

### Supplementary Note 2 The continuous model and its discretization

The epidemic model form [19, 1] (to which we refer for a precise derivation) provides a quantification of the number $Y(t,\tau,\tau')$ of people at time $t$ that have been infected at time $t-\tau$ by people who have in turn been infected at time $t-\tau'$.

The model characterizes $Y$ as a function of $s(\tau)$ and $\beta(\tau)$ (see Supplementary Note 1). Observe that both are quantities in $[0,1]$, and that $s(\tau)$ is non decreasing. The model then states that $Y(t,0,t)$ is a given initial value and that for $0 \leq \tau < t$ it holds

$$Y(t,0,\tau) = \beta(\tau)\left(1-\varepsilon_I s(\tau)\right)\int_\tau^t \left(1-\varepsilon_T \frac{s(\tau')-s(\tau'-\tau)}{1-s(\tau'-\tau)}\right) Y(t,\tau,\tau')d\tau'. \qquad (8)$$

The values of $\varepsilon_I, \varepsilon_T \in [0,1]$ are fixed in the original model, while we assume from now on that may depend on $\tau$. This dependence on the time is anyhow not used in the scenarios considered in this paper.

Observe that in the absence of containment policies (i.e. $\varepsilon_I = \varepsilon_T = 0$) the model predicts a behavior

$$Y(t,0,\tau) = \beta(\tau)\int_\tau^t Y(t,\tau,\tau')d\tau',$$

i.e. the new infected individuals are just given by the cumulative number of people who have been infected at previous times, weighted by the infectiousness of the disease. In other words, every previously infected person is a possible agent of new infection, and in this scenario an exponential growth is observed. The isolation and tracing measures, on the other hand, act as discounts on the number of available spreaders of the epidemic.

#### Supplementary Note 2.1 A more convenient form of the equations

As mentioned before, the model was analyzed in Fraser et al. [19], Ferretti et al. [1] by considering its asymptotic behavior as $t$ grows to infinity. We instead need a finite-time model that allows a flexible treatment of real data. To this end, it is convenient to use the variable $\Lambda(t,\tau) := Y(t,0,\tau)$ (as in Fraser et al. [19]) which represents the number of people which are infected at time $t$ by people who have been infected for time $\tau' \leq t$.

With straightforward manipulations, equation (8) can be rewritten for $0 \leq \tau < t$ as follows

$$
\begin{aligned}
Y(t,0,\tau) &= \beta(\tau)\left(1-\varepsilon_I(\tau)s(\tau)\right)\int_{\tau}^{t}\left(1-\varepsilon_T(\tau)\frac{s(\tau')-s(\tau'-\tau)}{1-s(\tau'-\tau)}\right)Y(t,\tau,\tau')d\tau' \\
&= \beta(\tau)\left(1-\varepsilon_I(\tau)s(\tau)\right)\int_{0}^{t-\tau}\left(1-\varepsilon_T(\tau)\frac{s(\rho+\tau)-s(\rho)}{1-s(\rho)}\right)Y(t,\tau,\rho+\tau)d\rho \\
&= \beta(\tau)\left(1-\varepsilon_I(\tau)s(\tau)\right)\int_{0}^{t-\tau}\left(1-\varepsilon_T(\tau)\frac{s(\rho+\tau)-s(\rho)}{1-s(\rho)}\right)Y(t-\tau,0,\rho)d\rho,
\end{aligned}
$$

where we changed the integration variable to $\rho := \tau' - \tau$, and we used the translational invariance of $Y$. In the variable $\Lambda$, this reads as

$$
\Lambda(t,\tau) = \beta(\tau)\left(1-\varepsilon_I(\tau)s(\tau)\right)\int_{0}^{t-\tau}\left(1-\varepsilon_T(\tau)\frac{s(\rho+\tau)-s(\rho)}{1-s(\rho)}\right)\Lambda(t-\tau,\rho)d\rho. \quad (9)
$$

Observe that this is an evolution equation that requires to define an initial number of infected individuals, i.e. we assume that the quantity $\Lambda(0,0) := \Lambda_0$ is a given number.

The quantity of interest is then the total number $\lambda(t) := \int_0^t \Lambda(t,\tau)d\tau$ of newly infected individuals at time $t$.

#### Supplementary Note 2.2 Discretization

We fix a value $T > 0$ as the maximal simulation time and take $n+1$ points in $[0,T]$ i.e., $\tau_i := i\left(\frac{T}{n}\right)$, $0 \le i \le n$.

We will approximate the values of $\Lambda(\tau_k,\tau_i)$ for $k = 1,\dots,n$ and $i = 0,\dots,k-1$, while, according to Fraser et al. [19], we set $\Lambda(\tau_k,\tau_i) = 0$ for all $i \ge k$. Moreover, we assume that the value $\Lambda(\tau_1,\tau_0)$ is given.

Observe that this discretization is equivalent to assume that the number of new cases is measured only at equal discrete times (e.g. at the end of each day) rather than measured continuously.

We show in the next section that the continuous model (9) can be approximated by defining a suitable value for $\Lambda(\tau_1,\tau_0)$, and then iteratively computing the values of $\Lambda(\tau_k,\tau_i)$ by applying the simple formula

$$
\Lambda(\tau_k,\tau_i) = \frac{T}{n}\sum_{j=0}^{k-i-1}\left(A_{\varepsilon_I,\varepsilon_T}\right)_{ij}\Lambda(\tau_{k-i},\tau_j), \quad 0 \le i < k \le n,
$$

where the matrix $A_{\varepsilon_I,\varepsilon_T} \in \mathbb{R}^{n\times n}$ is defined for $0 \le i,j \le n-1$ as

$$
\left(A_{\varepsilon_I,\varepsilon_T}\right)_{ij} := \begin{cases} \beta(\tau_i)\left(1-\varepsilon_I(\tau_i)s(\tau_i)\right)\left(1-\varepsilon_T(\tau_j)\frac{s(\tau_{j+i})-s(\tau_j)}{1-s(\tau_j)}\right) & \text{if } j \le n-i-1, \\ 0 & \text{if } j > n-i-1, \end{cases},
$$

We remark that this equation is a forward-in-time system, meaning that the computation of the values of $\Lambda(\tau, t)$ is obtained using only values of $\Lambda$ for previous time steps, which have thus already been computed. This is in contrast with the case of Fraser et al. [19] and Ferretti et al. [1], where an eigenvalue equation has to be solved, and only the asymptotic state can be estimated.

Moreover, we can use $\Lambda$ to compute

$$\lambda(\tau_k) = \sum_{i=0}^{k-1} \Lambda(\tau_k, \tau_i), \quad 1 \leq k \leq n. \tag{10}$$

We remark that equation (1) in the main text uses versions of $\varepsilon_I, \varepsilon_T$ that are constant in time.

#### Supplementary Note 2.3 Derivation of the discretization

As said above, we fix a value $T > 0$ as the maximal simulation time and take $n+1$ points in $[0, T]$ i.e. $\tau_i := i\left(\frac{T}{n}\right)$, $0 \leq i \leq n$.

The points will be used also to approximate integrals via a right-rectangle quadrature rule, i.e.

$$\int_0^{\tau_i} f(\tau) d\tau \approx \frac{T}{n} \sum_{j=0}^{i-1} f(\tau_j), \quad 1 \leq i \leq n. \tag{11}$$

The goal is to approximate the values of $\Lambda(\tau_k, \tau_i)$ for $k = 1, \ldots, n$ and $i = 0, \ldots, k-1$, while, according to Fraser et al. [19], we set $\Lambda(\tau_k, \tau_i) = 0$ for all $i \geq k$. Moreover, we assume that the value $\Lambda(\tau_1, \tau_0)$ is given.

For $1 \leq k \leq n$ we first evaluate (9) at the points, first in the variable $t$ for $1 \leq k \leq n$, i.e.

$$\Lambda(\tau_k, \tau) = \beta(\tau)\left(1 - \varepsilon_I(\tau)s(\tau)\right) \int_0^{\tau_k - \tau} \left(1 - \varepsilon_T(\rho)\frac{s(\rho+\tau) - s(\rho)}{1 - s(\rho)}\right) \Lambda(\tau_k - \tau, \rho) d\rho,$$

and then in the variable $\tau$ for $\tau < t$, that is for $0 \leq i < k \leq n$, i.e.

$$\Lambda(\tau_k, \tau_i) = \beta(\tau_i)\left(1 - \varepsilon_I(\tau_i)s(\tau_i)\right) \int_0^{\tau_k - \tau_i} \left(1 - \varepsilon_T(\rho)\frac{s(\rho+\tau_i) - s(\rho)}{1 - s(\rho)}\right) \Lambda(\tau_k - \tau_i, \rho) d\rho.$$

Now observe that for $0 \leq i < k \leq n$ we have

$$\tau_k - \tau_i = T\left(\frac{k}{n}\right) - T\left(\frac{i}{n}\right) = T\left(\frac{k-i}{n}\right) = \tau_{k-i},$$

which ranges between $\tau_k$ for $i = 0$ and $\tau_1$ for $i = k - 1$. The last equation becomes for $0 \le i < k \le n$

$$\Lambda(\tau_k, \tau_i) = \beta(\tau_i) \left(1 - \varepsilon_I(\tau_i) s(\tau_i)\right) \int_0^{\tau_{k-i}} \left(1 - \varepsilon_T(\rho) \frac{s(\rho + \tau_i) - s(\rho)}{1 - s(\rho)}\right) \Lambda(\tau_{k-i}, \rho) d\rho.$$

We can then use the quadrature rule (11) to discretize the integral and to obtain

$$\Lambda(\tau_k, \tau_i) = \beta(\tau_i) \left(1 - \varepsilon_I(\tau_i) s(\tau_i)\right) \frac{T}{n} \sum_{j=0}^{k-i-1} \left(1 - \varepsilon_T(\tau_j) \frac{s(\tau_j + \tau_i) - s(\tau_j)}{1 - s(\tau_j)}\right) \Lambda(\tau_{k-i}, \tau_j).$$

Observe that the upper limit in the sum has a value $0 \le k - i - 1 \le k - 1$ for $0 \le i < k$. Moreover, in this case we have for $0 \le j \le k - i - 1$ that

$$\tau_j + \tau_i = T\left(\frac{j}{n}\right) + T\left(\frac{i}{n}\right) = T\left(\frac{j+i}{n}\right) = \tau_{j+i},$$

which ranges between $\tau_i$ and $\tau_{k-1}$. Inserting this into the last equation we get for $0 \le i < k \le n$

$$\begin{aligned}\Lambda(\tau_k, \tau_i) &= \beta(\tau_i) \left(1 - \varepsilon_I(\tau_i) s(\tau_i)\right) \frac{T}{n} \sum_{j=0}^{k-i-1} \left(1 - \varepsilon_T(\tau_j) \frac{s(\tau_{j+i}) - s(\tau_j)}{1 - s(\tau_j)}\right) \Lambda(\tau_{k-i}, \tau_j) \\ &= \frac{T}{n} \sum_{j=0}^{k-i-1} \beta(\tau_i) \left(1 - \varepsilon_I(\tau_i) s(\tau_i)\right) \left(1 - \varepsilon_T(\tau_j) \frac{s(\tau_{j+i}) - s(\tau_j)}{1 - s(\tau_j)}\right) \Lambda(\tau_{k-i}, \tau_j). \end{aligned} \quad (12)$$

We can define the matrix $A_{\varepsilon_I, \varepsilon_T} \in \mathbb{R}^{n \times n}$ whose entries are defined for $0 \le i, j \le n - 1$ as

$$\left(A_{\varepsilon_I, \varepsilon_T}\right)_{ij} := \begin{cases} \beta(\tau_i) \left(1 - \varepsilon_I(\tau_i) s(\tau_i)\right) \left(1 - \varepsilon_T(\tau_j) \frac{s(\tau_{j+i}) - s(\tau_j)}{1 - s(\tau_j)}\right) & \text{if } j \le n - i - 1, \\ 0 & \text{if } j > n - i - 1, \end{cases},$$

which has a triangular structure (the first row is nonzero, in the second row the last element is zero, ..., in the last row only the first element is nonzero).

With this matrix we can rewrite (12) as

$$\Lambda(\tau_k, \tau_i) = \frac{T}{n} \sum_{j=0}^{k-i-1} \left(A_{\varepsilon_I, \varepsilon_T}\right)_{ij} \Lambda(\tau_{k-i}, \tau_j), \quad 0 \le i < k \le n, \quad (13)$$

which is a recursive equation that determines the evolution of $\Lambda(t, \tau)$ once an initial condition is given.

Assuming for now that these initial conditions are given, we can compute $\Lambda(\tau_k, \tau_i)$ forward in $k$ and backward in $i$. That is, after we computed $\Lambda(\tau_\ell, \tau_i)$ for all $\ell = 1, \ldots, k-1$, and for $0 \leq i < \ell$, we can use (13) to compute $\Lambda(\tau_k, \tau_i)$ for $1 \leq i < k$, since in this case the right hand side contains values $\Lambda(\tau_{k-i}, \tau_j)$ which have already been computed since $1 \leq k - i \leq k - 1$ for $1 \leq i < k$.

The only remaining case is $i = 0$, and in this case the formula (13) gives instead

$$\begin{aligned}\Lambda(\tau_k, \tau_0) &= \frac{T}{n} \sum_{j=0}^{k-1} \left(A_{\varepsilon_I, \varepsilon_T}\right)_{0j} \Lambda(\tau_k, \tau_j) \\ &= \frac{T}{n} \left(A_{\varepsilon_I, \varepsilon_T}\right)_{00} \Lambda(\tau_k, \tau_0) + \frac{T}{n} \sum_{j=1}^{k-1} \left(A_{\varepsilon_I, \varepsilon_T}\right)_{0j} \Lambda(\tau_k, \tau_j)\end{aligned}$$

thus

$$\Lambda(\tau_k, \tau_0) = \left(1 - \frac{T}{n} \left(A_{\varepsilon_I, \varepsilon_T}\right)_{00}\right)^{-1} \frac{T}{n} \sum_{j=1}^{k-1} \left(A_{\varepsilon_I, \varepsilon_T}\right)_{0j} \Lambda(\tau_k, \tau_j),$$

where

$$\begin{aligned}\left(A_{\varepsilon_I, \varepsilon_T}\right)_{00} &= \beta(\tau_0) \left(1 - \varepsilon_I(\tau_0) s(\tau_0)\right) \left(1 - \varepsilon_T(\tau_0) \frac{s(\tau_0) - s(\tau_0)}{1 - s(\tau_0)}\right) \\ &= \beta(\tau_0) \left(1 - \varepsilon_I(\tau_0) s(\tau_0)\right),\end{aligned}$$

and thus

$$\begin{aligned}\left(1 - \frac{T}{n} \left(A_{\varepsilon_I, \varepsilon_T}\right)_{00}\right)^{-1} \frac{T}{n} &= \frac{T}{n - T \left(A_{\varepsilon_I, \varepsilon_T}\right)_{00}} \\ &= \frac{T}{n - T\beta(\tau_0) \left(1 - \varepsilon_I(\tau_0) s(\tau_0)\right)}.\end{aligned}$$

This term is positive if and only if

$$0 < n - T\beta(\tau_0) \left(1 - \varepsilon_I(\tau_0) s(\tau_0)\right) \Rightarrow \beta(\tau_0) \left(1 - \varepsilon_I(\tau_0) s(\tau_0)\right) < n/T.$$

Since the left hand side is at most $\beta(\tau_0)$, it is sufficient to require that $n/T > \beta(\tau_0)$, or $n > \beta(\tau_0) \cdot T$.

In this way we defined $\Lambda(\tau_k, \tau_i)$ for all values $1 \leq k \leq n$ and $0 \leq i < k$. It remains to assign the value $\Lambda(\tau_1, \tau_0)$, which can be fixed to the initial value $\Lambda_0$.

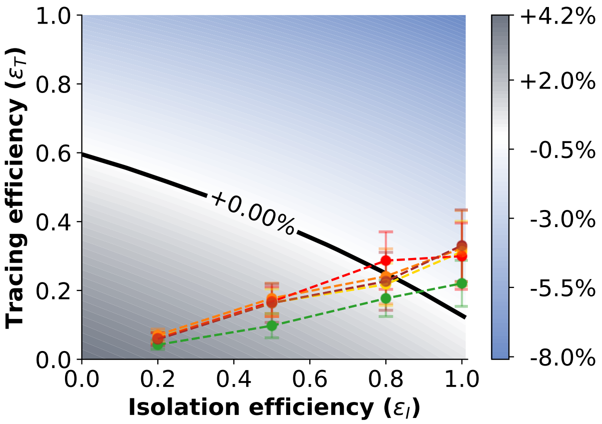


(a)

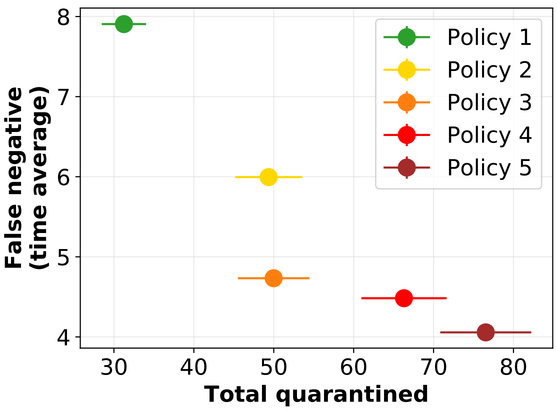


(b)

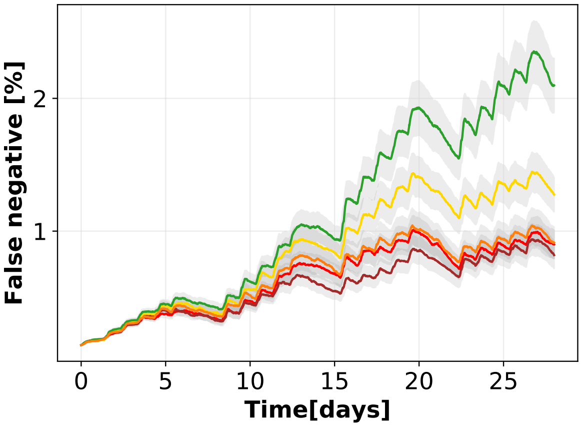


(c)

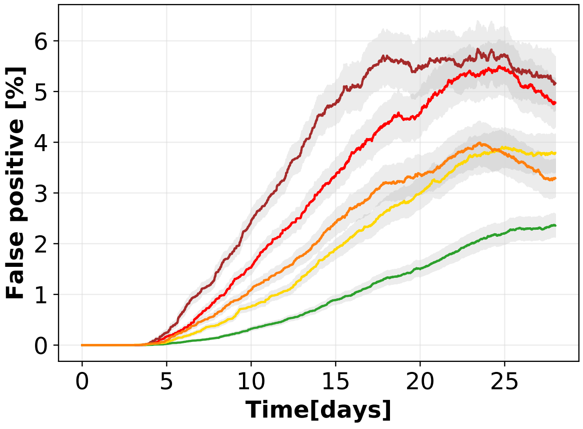


(d)

Supplementary Figure 6: **Tracing policy efficiency with longer contact memory: 15 (instead of 7) days.** 6a: Growth or decrease rate of the number of newly infected individuals and efficiency of the containment policies. 6b: Cross plot of the cost (number of quarantines) versus the effectiveness (low number of false negatives) for each policy. 6c and 6d: Temporal evolution of respectively the percentages of false negatives, i.e. infected individuals not quarantined, and false positives, i.e. not infected individuals quarantined, over the entire population, assuming an isolation efficiency of $\varepsilon_I = 0.8$, a reproductive number $R_0 = 1.5$, and 40% app adoption. The points in the first two panels and the curves in the last two have been obtained as mean values over 200 independent simulations, the corresponding error bars and the curve shadings represent the standard error.

### Supplementary Note 3 Evaluation of additional containment measures and refined policies

#### Supplementary Note 3.1 Longer and shorter tracing memory

We explore here how the outcomes of the different policies depend on the memory length of the contact history, which has been set to 7 days in the previous simulations (see Supplementary Notes of the main text).

First, to understand whether or not an increased memory would improve the effectiveness of each policy, we repeat the experiments assuming that the contacts of each individual are recorded for 15 days in the past, and report the results in Supplementary Fig. 6. When comparing Supplementary Fig. 6a with the original setting (central panel of Fig. 5 in the main text), it is clear that the increased memory brings a negligible advantage. This is confirmed by the total number of false negatives in Supplementary Fig. 6b if compared with Fig. 6c of the main text, and this is at the price of increased storage requirements, see total quarantines.

Second, it is worth investigating if a shorter tracing memory would give improvements in terms of the numbers of false positives. We thus repeat the simulations assuming that the memory is reduced to 2 days (still including the 2 days delay in the case reporting as in all other settings). Supplementary Fig. 7 shows that the shorter memory reduces the effectiveness of the policies of a significant amount, none of them crossing the black line for $\varepsilon_I = 0.8$. Apparently, storing only 2 days of contacts reduces too much the number of quarantined individuals (see Supplementary Fig. 7b), affecting the effectiveness.

#### Supplementary Note 3.2 Longer delay

The implemented model, for the sake of realism, includes a variable delay between the instant when a person is recognized as infected and the instant when that person is isolated. We set the delay to 2 days in all the other simulations and we test here the effect of a longer delay: 3 days, which is a good estimate for a system which is over-burdened but not close to collapse. From Supplementary Fig. 8a we observe that even one additional day of delay has a strong impact on the behavior of the epidemic, with none of the proposed policies able to cross the threshold of controllability, even for maximal isolation efficiency. Moreover Supplementary Fig. 8b shows that high levels of false negatives are reached for each policy, around twice those obtained with only two days of delay (see Fig. 6c in the main text) even if the total number of people in quarantine is slightly higher.

This highlights how rapid interventions are fundamental in containment policies based on contact tracing.

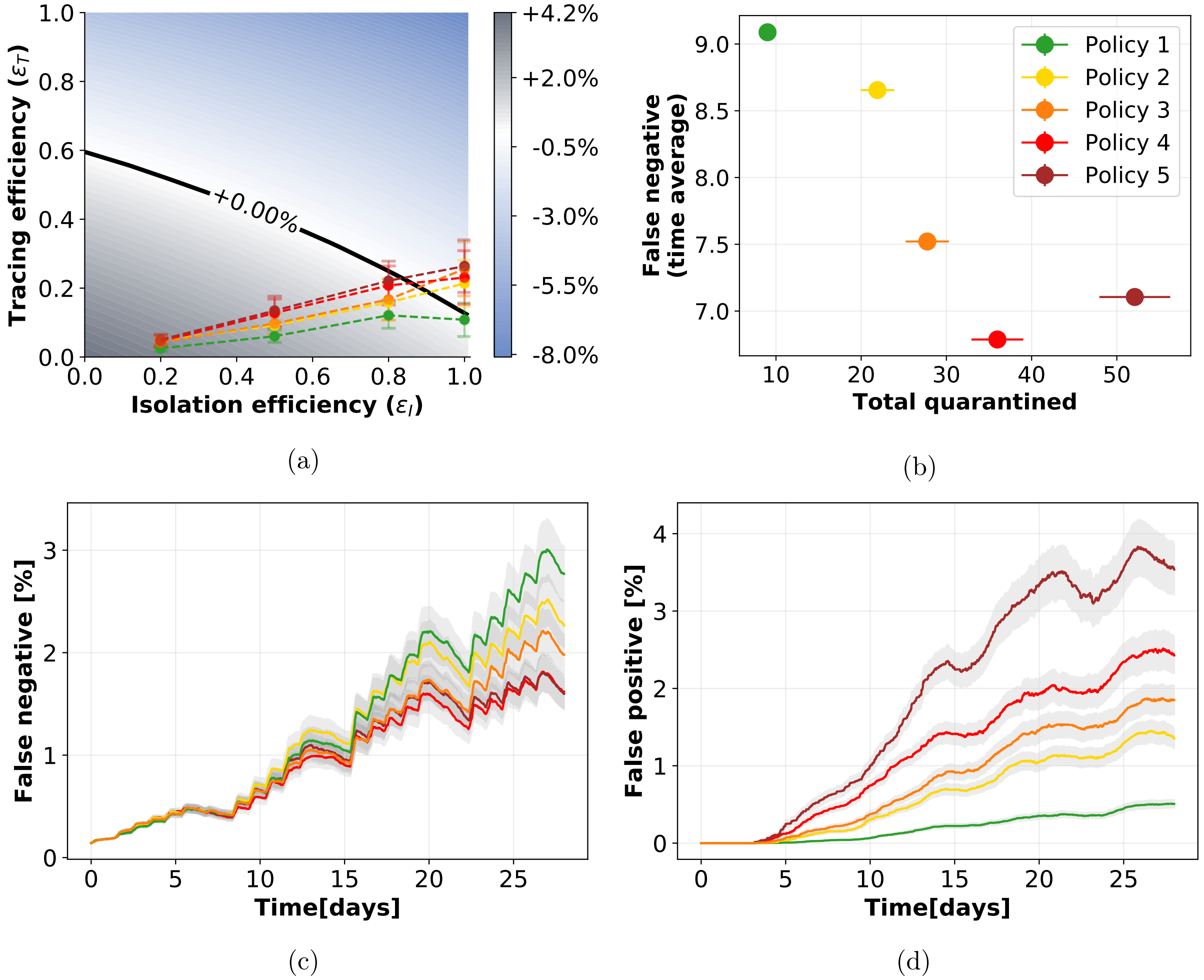


Supplementary Figure 7: **Tracing policy efficiency with shorter contact memory: 2 (instead of 7) days.** 7a: Growth or decrease rate of the number of newly infected individuals and efficiency of the containment policies. 6b: Cross plot of the cost (number of quarantines) versus the effectiveness (low number of false negatives) for each policy. 7c and 7d: Temporal evolution of respectively the numbers of false negatives, i.e. infected individuals not quarantined, and false positives, i.e. not infected individuals quarantined, assuming an isolation efficiency of $\varepsilon_I = 0.8$, a reproductive number $R_0 = 1.5$, and 40% app adoption. The points in the first two panels and the curves in the last two have been obtained as mean values over 200 independent simulations, the corresponding error bars and the curve shadings represent the standard error.

#### Supplementary Note 3.3 Second order tracing

We additionally explore the possibility to keep track of contacts in a recursive way. Namely, when an individual is isolated, not only its contacts are quarantined, but also its contacts'

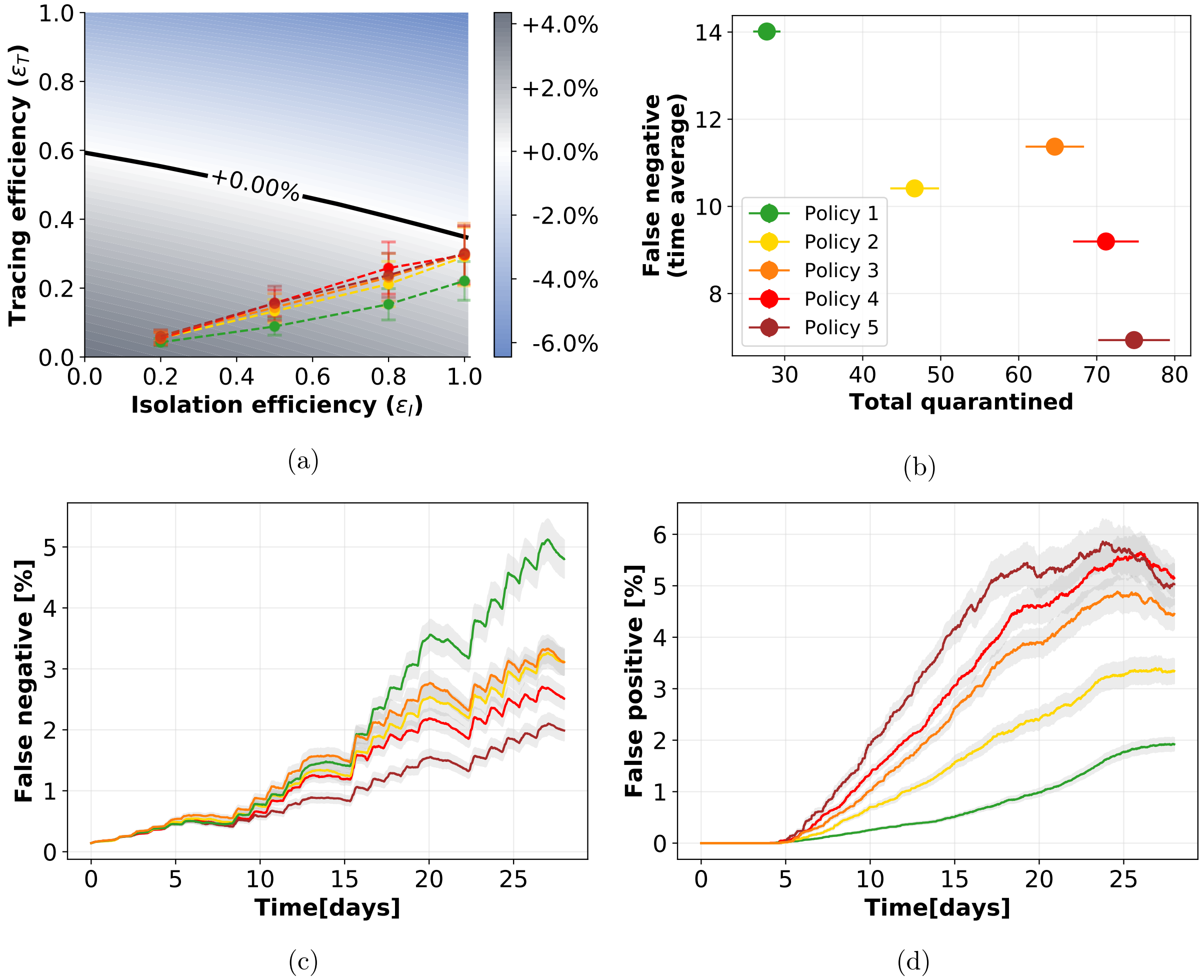


Supplementary Figure 8: **Tracing policy efficiency with a longer reporting delay: 3 (instead of 2) days.**
8a: Growth or decrease rate of the number of newly infected individuals and efficiency of the containment policies. 6b: Cross plot of the cost (number of quarantines) versus the effectiveness (low number of false negatives) for each policy. 8c and 8d: Temporal evolution of respectively the percentages of false negatives, i.e. infected individuals not quarantined, and false positives, i.e. not infected individuals quarantined, over the entire population, assuming an isolation efficiency of $\varepsilon_I = 0.8$, a reproductive number $R_0 = 1.5$, and 40% app adoption. The points in the first two panels and the curves in the last two have been obtained as mean values over 200 independent simulations, the corresponding error bars and the curve shadings represent the standard error.

contacts. This obviously means an enhanced risk in terms of preserving the privacy of individuals, and hence the major open question regarding this kind of policies is whether or not the increased intrusiveness into an individual's social network provides a tangible improvement of the virus containment efforts.

A complete study of this scenario is beyond the scope of this paper for a specific reason: the continuous model (see Supplementary Notes Supplementary Note 2) does not take into consideration this kind of tracing, and there is thus no way to use the information provided by the study of the data set in this framework.

Nevertheless, we find meaningful to report here the results of this additional experiment. We simulated the epidemic on the CNS data set, considering $R_0 = 1.5$, a delay of 2 days in isolating infected individuals and an app adoption of 40%. The numerical results are shown in Supplementary Fig. 9. We immediately notice that such intrusive tracing policy does not provide a significantly beneficial effect. Indeed, comparing Supplementary Fig. 9a and 9b with respectively Fig. 6a and 6b in the main text, which are the corresponding results for first order tracing, we notice that the levels reached by both false negatives and false positives are slightly reduced with second order tracing but not of a large amount. This appears clear also observing Supplementary Fig. 9c and the table, where the values of both total false negative and total quarantines are similar to those obtained with first order tracing (see Fig. 6c of the main text), with a slightly higher cost (larger percentages of quarantines) and a slightly larger effectiveness (lower false negatives).

This preliminary study seems to suggest that such a high level of tracing, which implies privacy issues (possibly even leading to lower adoption and compliance levels [20]), does not seem to be worth it since it is not going to provide meaningful improvements to the tracing system. We however remark once more that the reliability of this result is limited, being linked to a specific data set and not to a general theory. For this reason we observe that the concept of second-order tracing, a topic of recent discussions, deserves further investigation and may possibly be expanded in future works.

#### Supplementary Note 3.4 Variations in the number of asymptomatic individuals

In order to additionally verify the robustness of our predictions with respect to the epidemiological modelling, we assume here that the number of asymptomatic individuals is 20%, and additionally that a randomized testing policy that covers 25% of the asymptomatic population is in place.

In this case, little changes in the predictions of the model (Supplementary Fig. 10a) with respect to the case of 40% asymptomatics that was analyzed in the main text, since all the policies are effective for $\varepsilon_I = 1$, while Policy 1 is the only one that fails to contain the epidemic for $\varepsilon_I = 0.8$. No policy is effective for lower isolation efficiency. Similarly, the quarantine dynamics (false negative and false positive, Supplementary Fig. 10c and 10d) appear to have a similar behavior as in the basic setting. Despite these seemingly small changes in the success of the policies and in their cost, the cross visualization of Supplementary Fig. 10b shows that in this scenario it is harder to find a clear tradeoff between cost and effectiveness, since the two scores change smoothly between the five

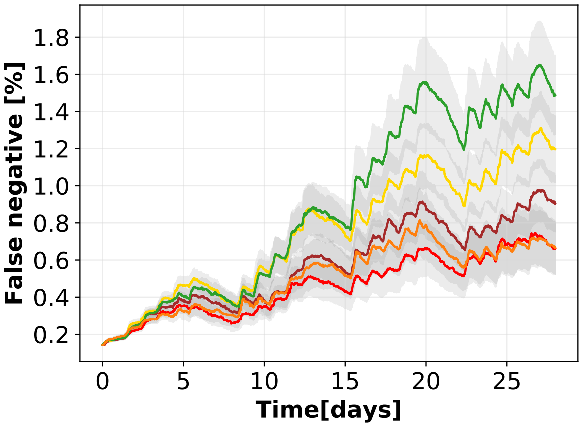


(a)

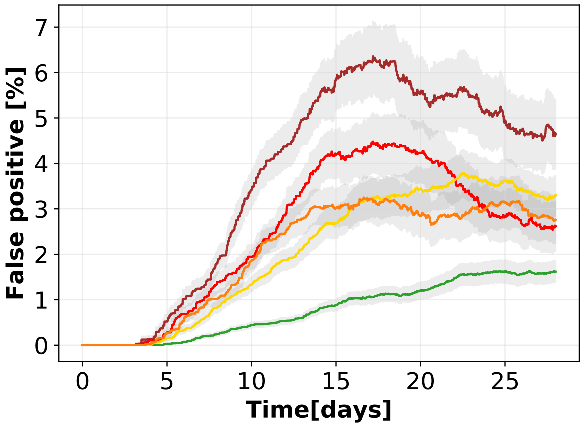


(b)

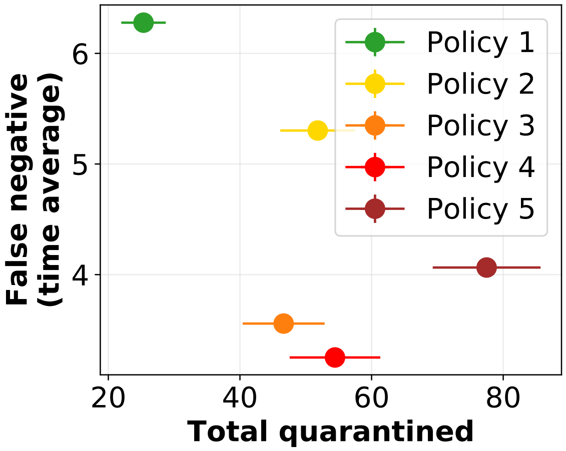


(c)

| Quarantined individuals | | |
|---|---|---|
| | % of quarantined over the entire population | % of infected over the quarantined population |
| ● | 4% | 10% |
| ● | 7% | 7% |
| ● | 7% | 6% |
| ● | 8% | 5% |
| ● | 11% | 4% |

Supplementary Figure 9: **Numerical simulations with second order tracing.** 9a and 9b: Temporal evolution of percentages of false negatives, i.e. infected individuals not quarantined, and false positives, i.e. not infected individuals quarantined, assuming an isolation efficiency of $\varepsilon_I = 0.8$. 9c: plot of the effectiveness (low number of false negatives) vs. cost (total quarantines) of the policies. The parameters are set so as to have $R_0 = 1.5$ and 40% app adoption. The table reports the percentage of distinct individuals who have been quarantined over the entire population and the percentage of them who were actually infected (true positive). The curves in the first two panels and the points in the third have been obtained as mean values over 100 independent simulations, the corresponding curve shadings and error bars represent the standard error.

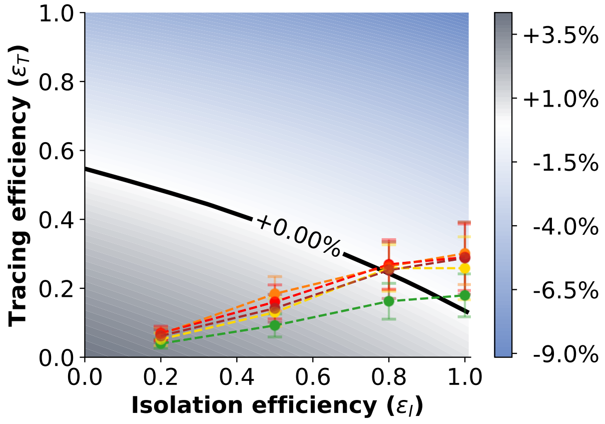


(a)

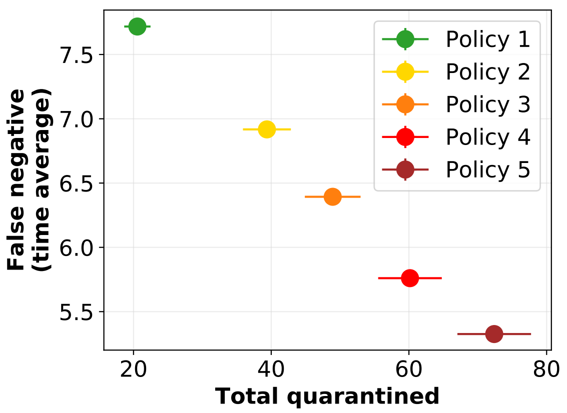


(b)

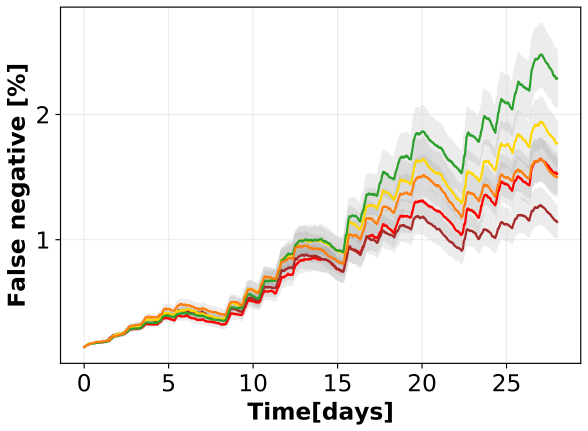


(c)

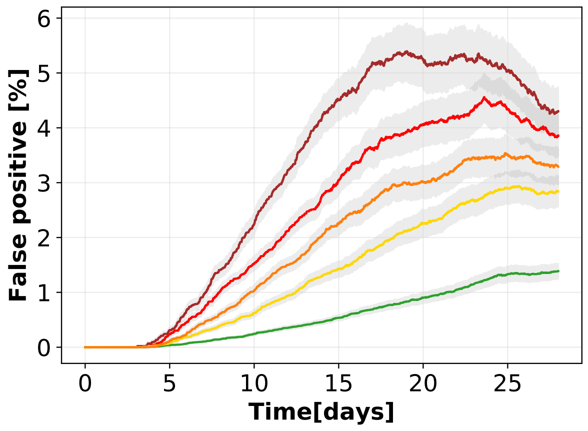


(d)

Supplementary Figure 10: **Tracing policy efficiency with 20% asymptomatic and 25% random testing.** 10a: Growth or decrease rate of the number of newly infected individuals and efficiency of the containment policies, assuming that symptomatic people account for the 80% of the infected individuals, that they can be isolated and that an additional 25% of asymptomatics can be identified via randomized testing. 10b: Cross plot of the cost (number of quarantines) versus the effectiveness (low number of false negatives) for each policy. 10d and 10d: Temporal evolution of respectively the percentages of false positives, i.e. not infected individuals quarantined, and false negatives, i.e. infected individuals not quarantined, over the entire population, assuming an isolation efficiency of $\varepsilon_I = 0.8$, a reproductive number $R_0 = 1.5$, and 40% app adoption. The points in the first two panels and the curves in the last two have been obtained as mean values over 200 independent simulations, the corresponding error bars and the curve shadings represent the standard error.

policies.

#### Supplementary Note 3.5 Close-range short-exposure vs long-range long-exposure interactions

We test here two additional policies obtained by mixing a low space resolution and a high time resolution, and viceversa. The policies are defined in Supplementary Fig. 12. Policy 6 delimits the risk to short exposure but close range interactions, while Policy 7 captures long exposure but long range interactions.

| ID | Signal strength (dBm) | Duration (min) | Fraction |
|---|---|---|---|
| ● Policy 6 | $-70$ | 5 | 17.9% |
| ● Policy 7 | $-91$ | 30 | 2.1% |

Supplementary Table 6: Parameters defining the two additional policies, and fraction of the total number of interactions of the CNS data set that they are able to detect.

Supplementary Fig. 11, in analogy with Supplementary Fig. 4 of the main text, shows the new policies overlaid to the histograms of duration and signal strength of the CNS data set contacts.

The values of the parameters $(\varepsilon_I, \varepsilon_T)$ characterizing the numerical simulations for the new policies with $R_0 = 1.5$ are shown in Supplementary Fig. 12a (see Fig. 5 in the main text, central panel, for a comparison with the policies in Fig. 3, main text), and it is clear that Policy 7 is as effective as the most restrictive policies (Policy 2 to Policy 5), while Policy 6 fails to contain the virus for an isolation efficiency smaller than 1. As for the policies of Fig. 3, this effectiveness comes at the cost of a larger number of quarantines (Supplementary Fig. 12c and Supplementary Fig. 12d). However, Supplementary Fig. 12b shows that the cost of Policy 7 is in larger than the ones of Policy 2 and Policy 3, but smaller than the ones of Policy 4 and Policy 5, while achieving a similar effectiveness.

We deduce that the ability to control the contagion seems to be more sensitive to duration of contacts than to their spatial distance. Indeed, policies which capture close range but short exposure interactions happen to be less performative in quarantining people than those signaling long range interactions with long exposure. In other words, quarantining individuals who have had a short interaction with an infected one, even if at close-range, is unnecessary. On the other hand, it appears to be important to track contacts with a high spatial resolution, including the ones that happens at a rather long distance, if their duration is significant.

However, we remark once more that these results are depending on the infectiousness model that we have defined here, and that they could possibly change in a different setting.

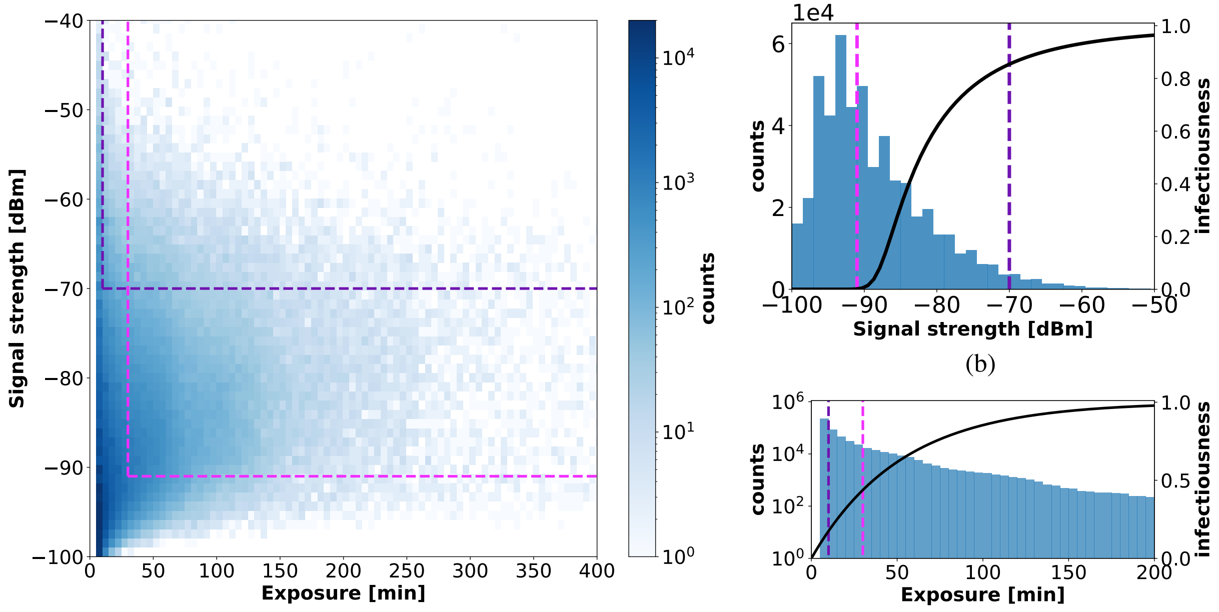


(a) (c)

Supplementary Figure 11: Distribution of the duration, panel (c), and signal strength (taken as a proxy for proximity), panel (b), of the contacts in the CNS data set. Panel (a) gives a scatterplot of signal strength vs duration, and displays the thresholds defining the two policies of Supplementary Table 6.

#### Supplementary Note 3.6 Compliance to quarantine decreases if notified multiple times

In the main text we consider compliance as encoding the compliance to all parts of the contact tracing and quarantine procedure. In other words, if some of the participants install the app but then do not quarantine if notified, then they should be counted among the non-compliant individuals since the effect would be the same than that of not adopting the app at all. The non-compliance (or impossibility) to quarantine is therefore already considered when choosing the percentage of app adoption. However, despite the fact that people who adopt the app are aware that they could be required to quarantine even if not infected, they may underestimate the possibility to be notified multiple times. A repeated quarantine could represent a relevant problem under social and economical aspects for many people, especially if unjustified. For this reason we decided to run an additional set of simulations where adoption of the app does not necessarily coincide with compliance to quarantine, and in particular it decreases if the same person is wrongly notified multiple times.

In particular we assume that compliance to quarantine can drop due to repeated notifications because the trust in healthcare and government institutions would drop too [21,

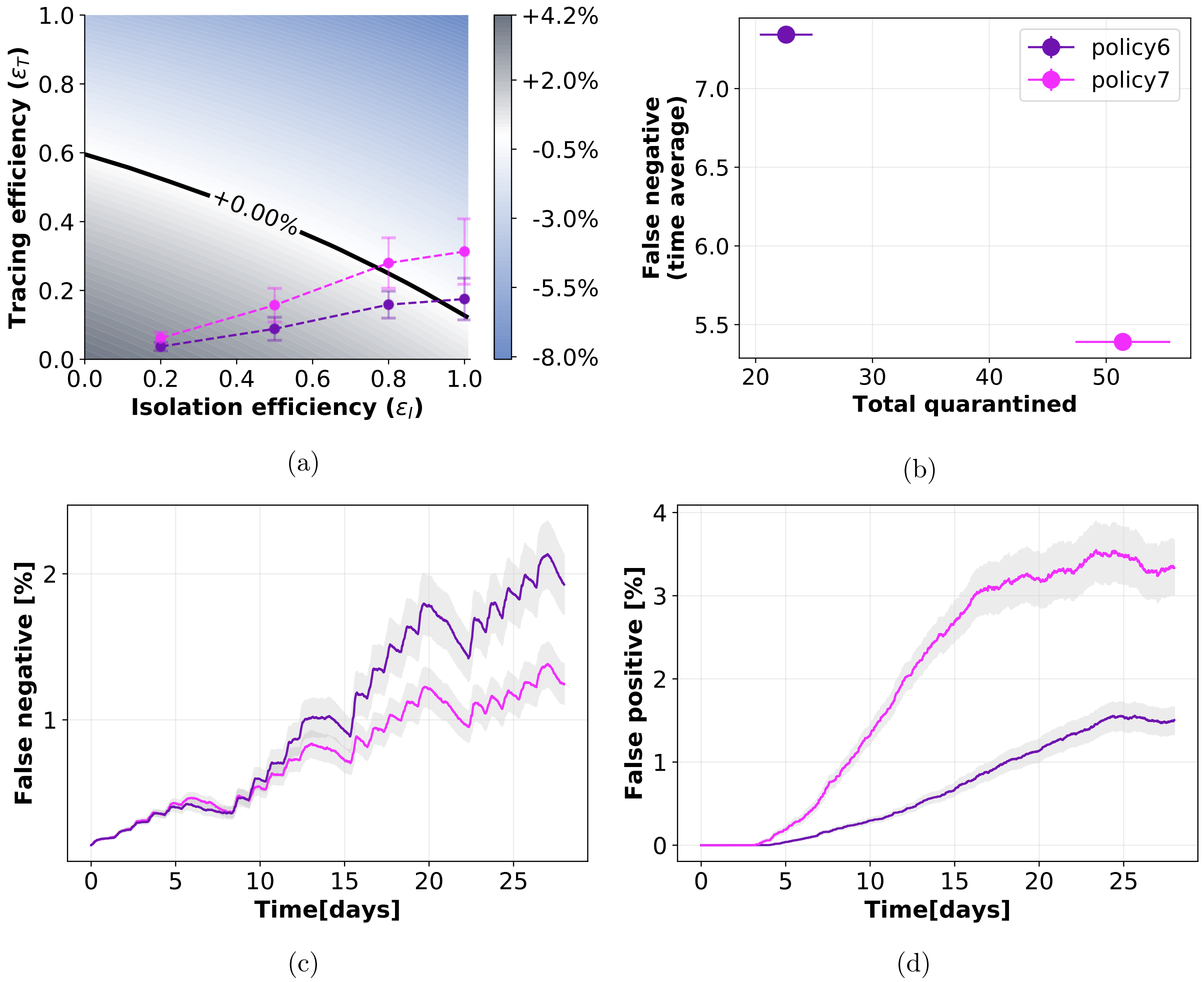


Supplementary Figure 12: **Tracing policy efficiency with additional policies.**
12a: Growth or decrease rate of the number of newly infected individuals and efficiency of the containment policies. 12b: Cross plot of the cost (number of quarantines) versus the effectiveness (low number of false negatives) for each policy. 12c and 12d: Temporal evolution of respectively the percentages of false positives, i.e. not infected individuals quarantined, and false negatives, i.e. infected individuals not quarantined, over the entire population, assuming an isolation efficiency of $\varepsilon_I = 0.8$, a reproductive number $R_0 = 1.5$, and 40% app adoption. The points in the first two panels and the curves in the last two have been obtained as mean values over 200 independent simulations, the corresponding error bars and the curve shadings represent the standard error.

22, 23]. Therefore the progressive decrease can be roughly estimated by considering the most classical game based on trust: the prisoner's dilemma [24, 25]. We focus in particular on an experiment of repeated game [26] where people were asked to play multiple rounds, each one with a different person. The experiment showed that willingness to cooperate decreased at each round and was measured for 10 rounds in total. We consider that the same reduction in trust can be applied to the willingness to quarantine if notified. In a broad sense, these two settings are indeed similar: in the prisoner's dilemma each person can choose to cooperate, which they know would be the best option for everybody, but they do it at their own expenses, while in alternative they can choose an egoistic strategy, putting the others at risk. In case of notification from the contact tracing app, people would undergo a sort of "quarantine dilemma". Indeed there are two possible choices: the compliant one (for the social benefit, but possibly in detriment of their own social and economic life) and the egoistic one where a person decides not to quarantine, putting at risk all the others.

We therefore consider that the first time that people are traced and identified as possible infected they quarantine with probability 1. The second time it happens, if the person did not develop symptoms during the first quarantine, the probability drops to 0.86. The third time to 0.6, and so on, according to the values in Supplementary Table 7.

| Previous quar. | 0 | 1 | 2 | 3 | 4 | 5 | 6 | 7 | 8 | 9 |
|---|---|---|---|---|---|---|---|---|---|---|
| Compliance | 1 | 0.86 | 0.60 | 0.57 | 0.49 | 0.46 | 0.43 | 0.41 | 0.40 | 0.29 |

Supplementary Table 7: The second row reports the probabilities of compliance to quarantine if notified by the app, given that the same person has already been quarantined, even if not infected, a number of times reported in the first row. The level of compliance have been chosen according to Ref. [26].

We simulated this setting on an extended version of the CNS data set, containing contacts for a period of three months instead of one, in order to be able to catch all the repeated notifications (see SI Supplementary Note 1.4 for a description of the extended time period).

Notice that this modification can be inserted into the mathematical model if we consider that the $\varepsilon_T$, that we compute as explained in Section  of the main text, changes its meaning. In this case it does not represent the ability to trace people but the possibility to quarantine them, since traced individuals could refuse to quarantine. Only for this case we thus rename $\varepsilon_T$ into $\varepsilon_Q$. The controllability of the epidemic is depicted in Supplementary Fig. 13a, while in Supplementary Fig. 13b we report the number of people who have been requested to quarantine as a function of the number of repetitions of these requests, for the five different policies. The time evolution of false negatives is depicted in Supplementary Fig. 13c. In general, in Supplementary Fig. 13 we observe a similar behavior to the one obtained in the original setting (Fig. 5 central panel and Fig. 6 in the main text), with a slightly general reduction of the efficacy of containment. Indeed, only few people are asked to quarantine multiple times, as shown by Supplementary Fig. 13b. We can therefore assume that the original setting that we chose – and used in all other simulations – depicts a scenario which is not far from the one that we obtain with this additional characteristic making the system more realistic, thus confirming the robustness of our model.

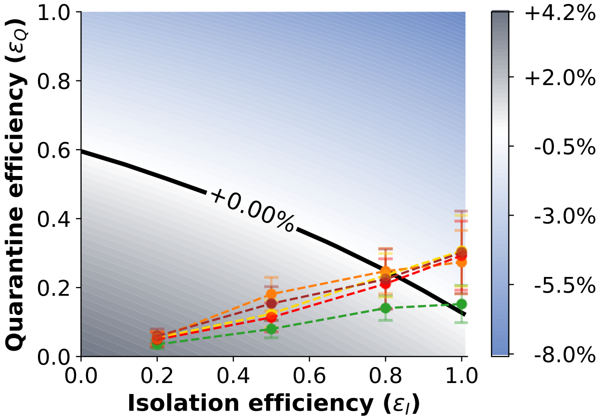


(a)

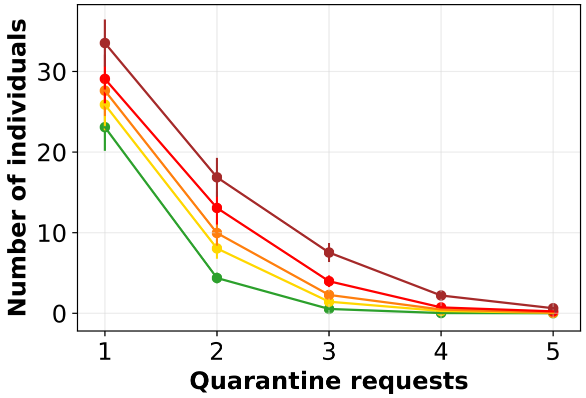


(b)

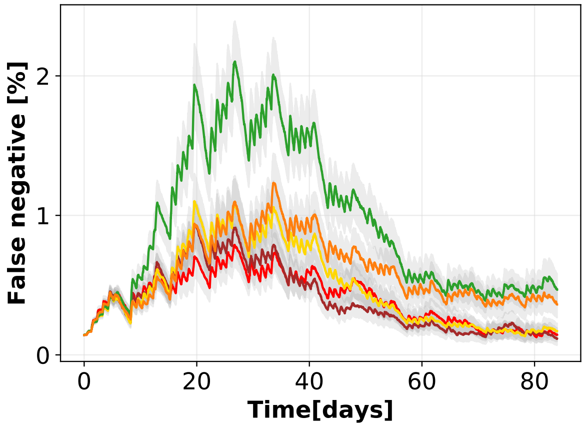


(c)

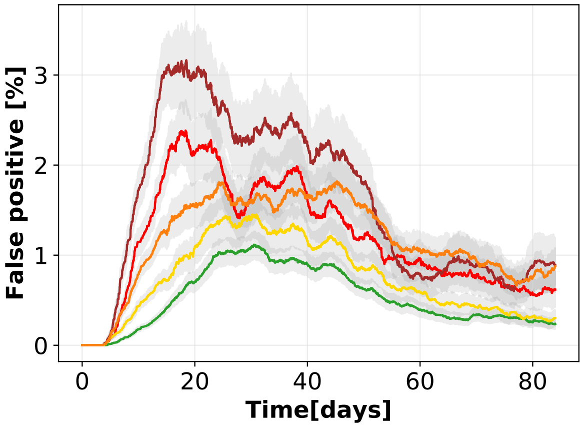


(d)

Supplementary Figure 13: **Compliance to quarantine variable in time.** 13a: Growth or decrease rate of the number of newly infected individuals and efficiency of the containment policies. 13b: Number of people who have been requested to quarantine as a function of the number of repetitions of these requests, for the five different policies. 13c and 13d: Temporal evolution of the percentages of respectively false negatives, i.e. infected individuals not quarantined, and false positives, i.e. not infected individuals quarantined, over the entire population, assuming an isolation efficiency of $\varepsilon_I = 0.8$, a reproductive number $R_0 = 1.5$, and 40% app adoption. The points in the first two panels and the curves in the last two have been obtained as mean values over $n = 100$ independent simulations, the corresponding error bars and the curve shadings represent the standard error.

The possibility to run the code on the extended data set provides in addition the possibility to observe the phenomenon of growth and decrease of the active infected, which after one month and a half dampen down, almost extinguishing the epidemic. The false negative peak is followed by the false positive and unjustified quarantines are reduced to almost zero in a couple of months (see Supplementary Fig. 13d).

### Supplementary Note 4 Contagion heterogeneity due to social structure

Each social context can be described by a temporal network of connections characterized by a complex and unique topology which reflects the structure and organization of the specific slice of society under study. Societies are usually organized in clustered structures and it is often possible to divide people in subgroups, or clusters of individuals who are more connected among each other than with individuals of other groups.

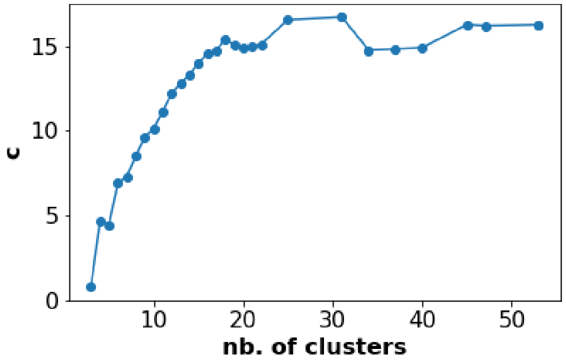


Supplementary Figure 14: **Intra- and inter-cluster contagion.** The ratio $c$, quantifying the tendency of contagions to take place inside a cluster rather than among different clusters, is reported for different possible choices of partitions corresponding to different numbers of clusters.

In this section we explore the clustering structure of the CNS data set and how contagion events are related to it. We use the Louvain algorithm for community detection [27], able to rapidly extract the community structure even for large networks. We apply it to the aggregated graph of CNS, which is obtained by transforming the temporal graph in a static one by considering at the same time all the connections among students and weighting them according to their intrinsic characteristics: duration and proximity. In particular, we define the aggregated graph by assigning to the edges with a proximity below the threshold of -90 dBm a unitary weight, while for all the contacts characterized by a closer proximity we label each edge with the total duration of contacts between the corresponding pair of nodes. By modifying the resolution of the algorithm we achieve different possible partitions of the students, corresponding to different numbers of clusters, from 3 to 53. We then simulate our model of contagion with isolation and tracing. Taking into account a sample of fifty simulations, we count how many contagions in average take place intra- and inter-cluster for each of the chosen partitions of the network. The numbers of contagions are normalized with the numbers of existing contacts respectively intra- and inter-cluster, and we define the ratio:

$$c = \frac{\text{fraction of intra-cluster contagions}}{\text{fraction of inter-cluster contagions}} \tag{14}$$

which quantifies the tendency of contagions to take place inside a cluster rather than among different clusters. For all the possible partitions based on duration of contacts we find $c > 1$, as reported in Supplementary Fig. 14.

We then observe the time evolution of the fraction of infected individuals belonging to each different cluster. In Supplementary Fig. 15 this is depicted for two different possible partitions and two different policies. In the first chosen partition the sample of students is divided into seven clusters. We observe that, even if the levels of infections are different for the two policies, being Policy 5 far more restrictive and thus effective than Policy 1, the curves present a similar behavior (see panels (a) and (b)). In particular, there is one group, identified by label 3, which is statistically more at risk, since it is observed that contagions diffuse faster than in the other groups. Let us point out that such cluster is not the most numerous one, as shown from the bottom left table in Supplementary Fig. 15. The rapid growth of the curve of infections in the “front-runner" cluster is immediately followed by four other groups, which evolve roughly together. The slower ones, with very few contagions, are the less numerous clusters, which have few contacts with the rest of the population.
The second partition that we take into account is composed by fifteen clusters. Again, we observe that different groups show similar behaviors with the exception of one group with a faster spreading and the small clusters, which tend to be preserved.

The study of contagions within and among socially connected clusters of individuals and how the general epidemic depend on the organization of the network in these substructures represents a rich and fascinating field of analysis [28]. Moreover, differentiated policies could take into account this particular social structure, as proposed by Block et al. [29]. Nevertheless a deeper analysis on this topic is out of the scope of the present manuscript and a future study will possibly be devoted to it.

### Supplementary Note 5 Extended results on SocioPatterns data sets

In this section we present the results of simulations performed on two different data sets: (i) HighSchool13[30], collected in a French high school, and (ii) InVS15[31], collected in a French workplace. Both data sets have been collected using the sensing platform developed by the SocioPatterns collaboration[1], based on wearable proximity sensors that exchange

[1]http://www.sociopatterns.org/

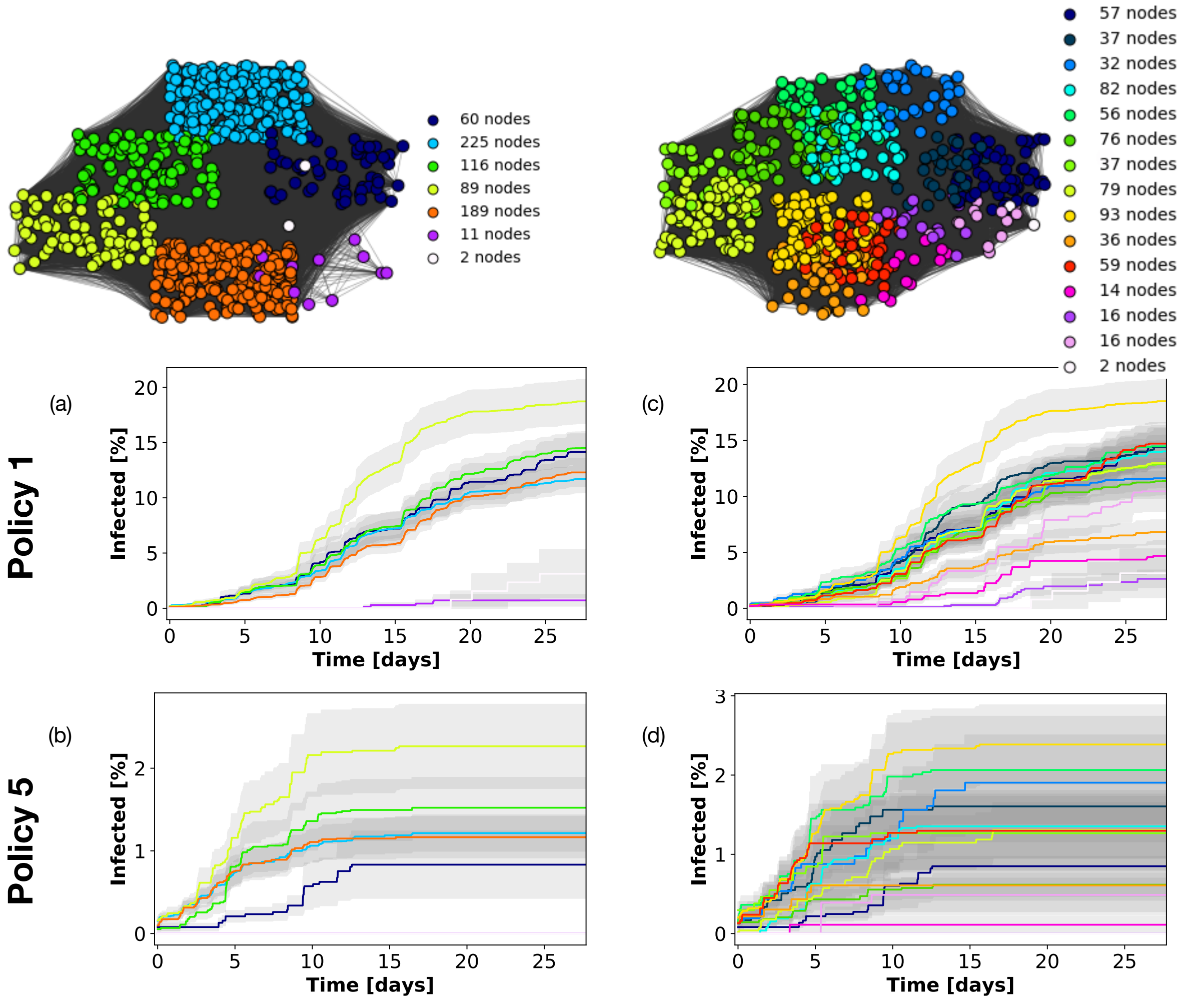


Supplementary Figure 15: **Infected individuals in network clusters** The two graphs above depict two possible partitions of the aggregated network of interactions for the CNS data set. The number of nodes in each cluster is reported in the legend. Panels (a) and (b) show the time evolution of the percentage of infected individuals in each of the seven clusters of the first partition, respectively applying Policy 1 and Policy 5. Panels (c) and (d) analogously represent infections in the fifteen clusters of the second partition, respectively for Policy 1 and Policy 5. The curves have been obtained as mean values over 100 independent simulations and the curve shadings represent the standard error.

radio packets, detecting close proximity ($\leq 1.5m$) of individuals wearing the devices [32]. These data do not contain information on the signal strength, but simply give a list of contacts between individuals with a resolution of 20 seconds. Both simulations and policies are thus redefined only as a function of contact durations.

In order to study the effectiveness of the policies and the spreading of the virus, and given the timescales involved, we need data extending on more than 15 days. As the SocioPatterns data have a high temporal resolution (20 seconds) but were collected for shorter overall durations, we artificially extend the length of each data set by replicating it (copying and pasting the entire data set so as to concatenate it multiple times). Supplementary Table 8 gives the number of nodes, the length of the data set (in days) and the duration of the replicated data.

| | InVS15 | HighSchool13 |
|---|---|---|
| # of nodes | 211 | 327 |
| Days | 11.5 | 4.2 |
| Extended Days | 46 | 16.8 |

Supplementary Table 8: Number of nodes, days and extended days for each SocioPatterns data set.

For both these data sets, similarly to the CNS data set, most contacts happen before the infectiousness reaches its peak (Supplementary Fig. 16), even if contacts are present for all possible durations. Nevertheless, these are sufficient to spread the infection.

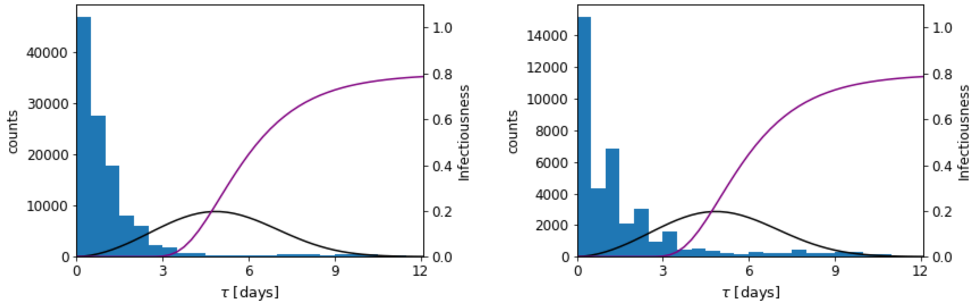


(a) (b)

Supplementary Figure 16: **Infectiousness and contact distribution in a high school and in an office.**
Distribution of the time since infection of the people having contacts, probability distribution $\omega(\tau)$ (black line) determining the infectiousness as a function of time, and distribution $s(\tau)$ determining the cumulative probability to detect an infected person (purple line). The two plots are obtained with $\varepsilon_I = 0.8$ and Policy 5 for the InVS15 (Supplementary Fig. 16a) and the HighSchool13 data sets (Supplementary Fig. 16b).

We further run the simulations on the network for the five policies of Fig. 3 in the main text (recall that only distances are taken into account). To this end, the scaling factors of the infectiousness (Supplementary Note 1.2) require first a calibration. We thus first recompute the factors $p_{R_0}$ to obtain $R_0 = 3$, and then we compute $r_{R_0}$ as reported in Supplementary Table 9.

| | InVS15 | HighSchool13 |
|---|---|---|
| $R_0$ | 1.5 | 1.5 |
| $r_{R_0}$ | 0.49 | 0.35 |
| $p_{R_0}$ | 420 | 252 |

Supplementary Table 9: Reduction factors for the SocioPatterns data sets.

#### Supplementary Note 5.1 SocioPatterns data: High School

In this case no policy is able to reach containment for $\varepsilon_I = 0.8$ (Supplementary Fig. 17a), even if Policy 5 is essentially on the boundary of no epidemic growth. For $\varepsilon_I = 1$, instead, all the policies are successful.

The uniform lack of success of the five policies is reflected by a similar time evolution of the curves of the number of false negatives (Supplementary Fig. 17c). Still, the policies are uneven regarding their quarantine cost (Supplementary Fig. 17d), since Policy 5 wrongly quarantines a substantially larger number of people than the other ones. In this case, the cost to effectiveness plot (Supplementary Fig. 17b) does not identify a best policy. This is to be expected, since none of them achieve the goal, and thus side costs play a little role in the ranking of the policies.

#### Supplementary Note 5.2 SocioPatterns data: Workplace

In the workplace environment, Policy 5 is successful for $\varepsilon_I = 0.8$ (Supplementary Fig. 18a), and for $\varepsilon_I = 1$ all policies except for Policy 1 are successful.

The higher effectivity of Policy 5 is reflected in a higher cost in terms of false positives (Supplementary Fig. 18d), but in this case this information is not particularly meaningful since there is no other successful policy to compare with. This is also the case of the cost to effectivity comparison (Supplementary Fig. 18b).

We also remark that in the numerical simulation the number of false negatives is in fact rapidly dropping to zero for all policies (Supplementary Fig. 18c), and this might suggest that the epidemic spread is kept under control. It is a peculiar case, since that the data set contains very little contacts, so the epidemic is spreading on few people (often 2-3) without propagating further. On the other hand, missing just one of the infected from tracing results into a very high ratio of unsuccessful tracing, thus in a small value of $\varepsilon_T$. This value, when inserted into the mathematical model, predicts the no-containment outcome observed before (Supplementary Fig. 18a). This seemingly contradictory behavior is in

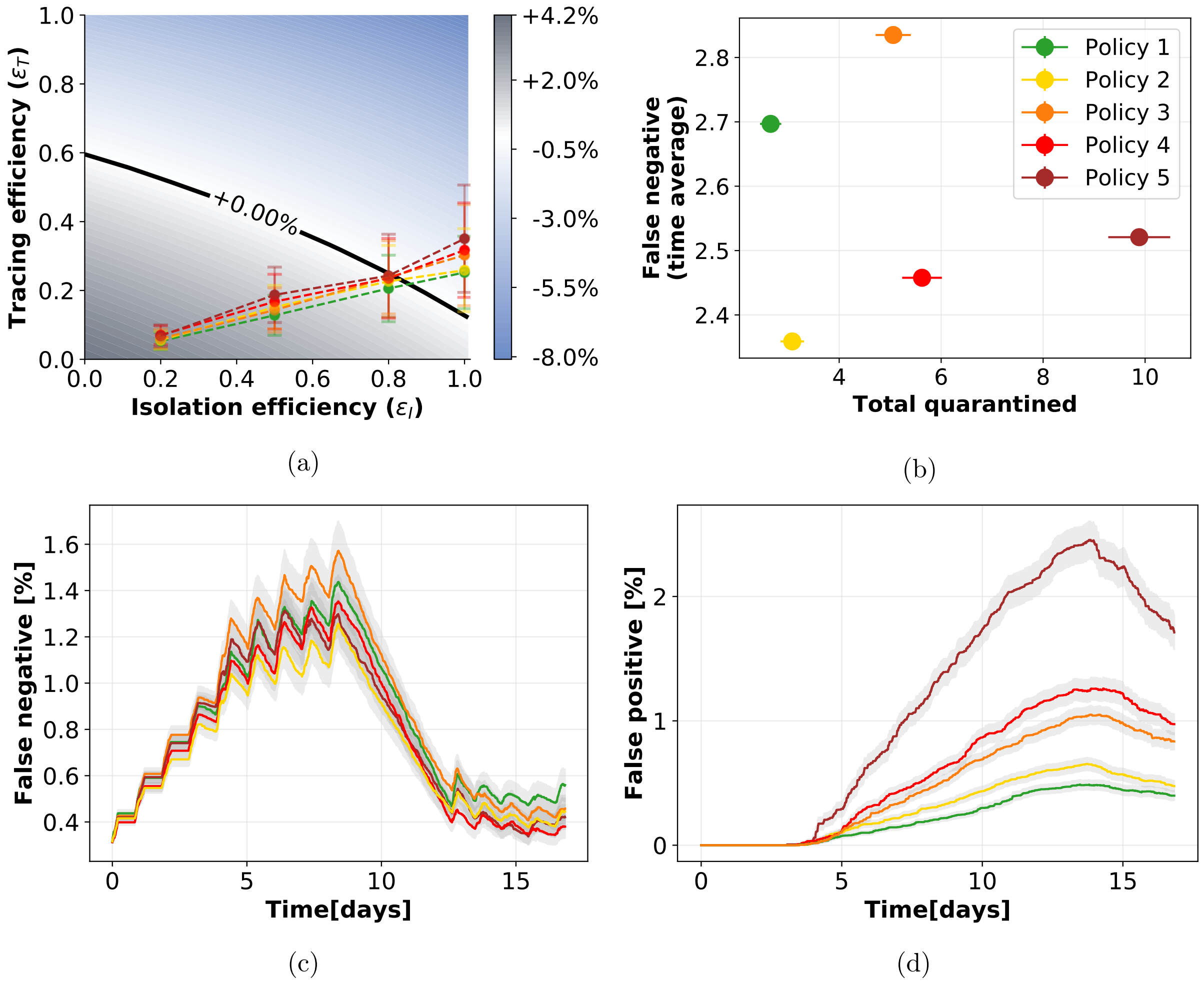


Supplementary Figure 17: **Tracing policy efficiency in a high school.** 17a: Growth or decrease rate of the number of newly infected individuals and efficiency of the containment policies. 17b: Cross plot of the cost (number of quarantines) versus the effectiveness (number of false negatives). 17c and 17d: Temporal evolution of respectively the percentages of false negatives, i.e. infected individuals not quarantined, and false positives, i.e. not infected individuals quarantined, over the entire population, assuming an isolation efficiency of $\varepsilon_I = 0.8$, a reproductive number $R_0 = 1.5$, and 40% app adoption. The points in the first two panels and the curves in the last two have been obtained as mean values over 200 independent simulations, the corresponding error bars and the curve shadings represent the standard error.

fact only revealing the fact that the mathematical model works on aggregated quantities, assuming homogeneous contacts, and in this case the spreading is a rare event in the network that is not possible to capture effectively by averaging over the entire population.

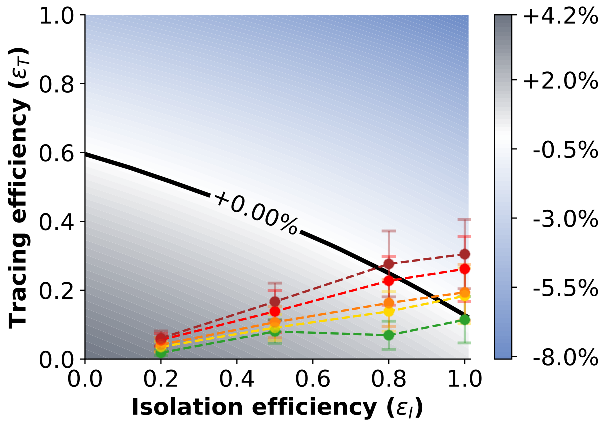


(a)

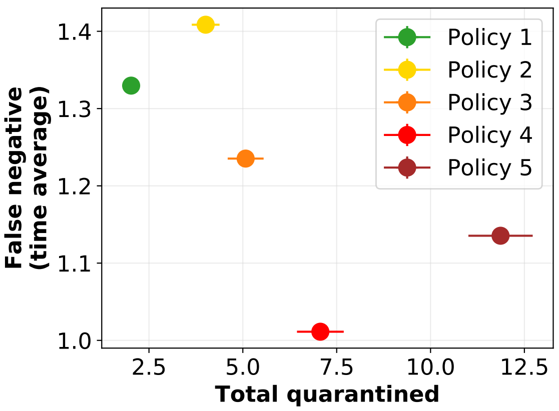


(b)

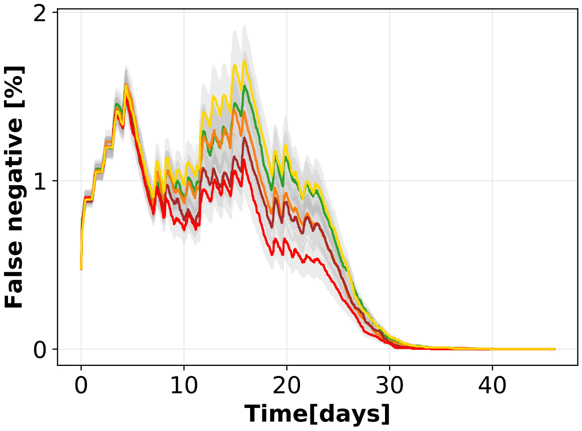


(c)

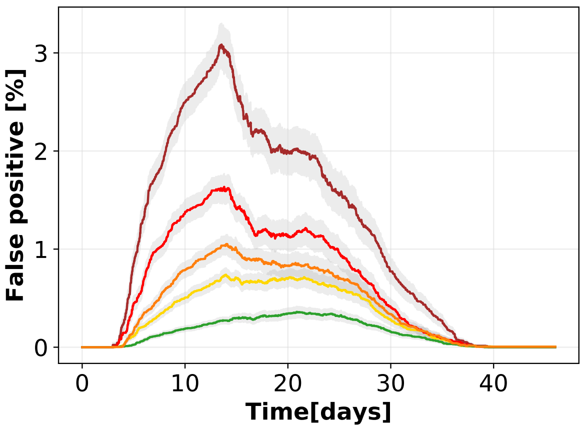


(d)

Supplementary Figure 18: **Tracing policy efficiency in an office building.** 18a: Growth or decrease rate of the number of newly infected individuals and efficiency of the containment policies. 18b: Cross plot of the cost (number of quarantines) versus the effectiveness (number of false negatives). 18c and 18d: Temporal evolution of respectively the percentages of false negatives, i.e. infected individuals not quarantined, and false positives, i.e. not infected individuals quarantined, over the entire population, assuming an isolation efficiency of $\varepsilon_I = 0.8$, a reproductive number $R_0 = 1.5$, and 40% app adoption. The points in the first two panels and the curves in the last two have been obtained as mean values over 200 independent simulations, the corresponding error bars and the curve shadings represent the standard error.

### Supplementary Note 6 Other models of digital contact tracing

Multiple recent modeling studies have shown that contact tracing may reduce epidemic spreading, and that the efficacy of its realization – contact identification and timing – plays a pivotal role for mitigation.

Some theoretical models of contact tracing date back to 2003-2004 and were originally developed for fighting smallpox [33, 34], proving that isolation and tracing are useful to slow down contagions. These works however have a slightly different approach where contact tracing is limited to small communities, which would be useless with a large-scale epidemic like that of COVID-19. Multiple alternative procedures have been proposed in the last months. For instance, the works of Hellewell et al. [35] and of Kretzschman et al. [36]. The first one assumes that contacts are traced with a fixed probability $\rho$, while the second one includes a distinction between contacts within a household and outside – these last not affected by physical distancing. Gorji et al. [37] instead propose a model where contact tracing is combined with a mass testing becoming "smart testing", suggesting that it would avoid numerous quarantines. A further work by Fraser et al. [38] explores the difference between centralized and decentralized tracing and the relative privacy issue. Backward and forward (predictive) tracing is introduced by Kojaku et al. [39], claiming that it could prevent a significant fraction of further transmissions. Very few studies make use of real-world contacts. A comprehensive study on isolation and tracing simulated on a real-world social network is provided by Firth et al. [40]. This study is however limited by the fact that the absence of targeted policies implies a large portion of the population being quarantined, with diffused local lockdowns. Refined policies are instead proposed by Lorch et al. [41], based on the risk of exposure of each individual in the specific sites they visit, making use of mobility data and crowding. A further numerical analysis is devised by Barrat et al. [42], applied to different social contexts. Another way to simulate spreading and tracing on realistic scenarios is represented by the use of data to generate synthetic contact networks. This approach has been devised by Kucharski et al. [43], Lopez et al. [44], Hinch et al. [45], and Abueg et al. [46]. Some of the works cited above are summarized in Ref. [47].

We claim that a complete analysis of contact tracing needs real or realistic data and at the same time should be based on a solid mathematical model. This model should be general enough in order to provide a framework that can be applied in multiple contexts. We also claim that specific and targeted policies should be implemented in order to control such a large-scale epidemic without implying a total or partial lockdown.

The mathematical framework that we chose to implement in our strategy, as previously mentioned, is that proposed by Fraser et al. [19] and its adaptation to the COVID-19 pandemic (Ferretti et al. [1]). This work describes the evolution of an epidemic in a homogeneously mixed population. It uses recursive equations that have been adapted to include the parameters $\varepsilon_I$ and $\varepsilon_T$ and, assuming an exponential growth for the number of infected individuals, the authors study how the growth rate depends on the intervention parameters. This inspiring approach represents the baseline of our model, which, not only enriches the work of Ferretti et al., but goes beyond the original analyses opening a wider scenario allowing policy evaluation. First, the assumption of full homogeneous mixing represents a limitation in epidemic modeling [48, 49, 10, 50], while realistic social network architectures might be particularly relevant for contact tracing [39]. We overcome this problem by obtaining contact tracing efficiency from numerical simulations on real-world contacts, thus capturing complex interaction structures that are necessary for a realistic quantification of this parameter. Second, in the work of Fraser et al. the mathematical framework is limited to exponential growth, and we devised a modified version of the equations where the time evolution is not constrained to any specific form. Third, we considered the parameter $\varepsilon_T$ to be dependent from $\varepsilon_I$, since tracing is a direct consequence of isolation: we would not have tracing without first identifying and isolate the primary cases. Moreover, for what concerns the epidemiological aspect, it is true that the infectiousness rate in the work of Fraser et al. is accurately designed based on literature and data, including contributions of asymptomatics too, however the symptom onset rate is defined such that everyone sooner or later gets symptoms. This implies that every individual can possibly be isolated, altering the reliability of the isolation procedure. We corrected this detail too, requiring that the curve in Supplementary Fig. 4 of the main text goes to 0.8 at large times. In general, we have slightly modified the epidemiological aspect of the model, using recent literature on COVID-19 [14, 51, 13], to consider asymptomatic cases and the delay in isolating individuals after they are identified as infected (Supplementary Note 2). Finally, as previously highlighted, the distinctive characteristic of our work is the evaluation of tracing efficiency on real contact data captured by Bluetooth sensors, and no more on an arbitrary parameter of the model. In particular, our work focuses on investigating how much the efficiency of DCT is influenced by the definition of different thresholds on the duration of exposure time and on the physical distance of detected contacts. This allows to devise appropriate policies and to evaluate which of them are more suitable after a study of efficiency and cost.

### Data Availability

The data that support the findings of this study are publicly available.
The CNS data can be found at `https://doi.org/10.6084/m9.figshare.7267433`
and the SocioPatterns data at `http://www.sociopatterns.org`

### Code Availability

We are pleased to make available the source-code accompanying this research [52]. The code uses Python (version 3.8.3), Numpy (version 1.18.5), Scipy (version 1.2.0), Networkx (version 2.5), Matplotlib (version 3.0.2).

### References

[1] Luca Ferretti, Chris Wymant, Michelle Kendall, Lele Zhao, Anel Nurtay, Lucie Abeler-Dörner, Michael Parker, David Bonsall, and Christophe Fraser. Quantifying SARS-CoV-2 transmission suggests epidemic control with digital contact tracing. *Science*, 2020.

[2] Yang Liu, Li-Meng Yan, Lagen Wan, Tian-Xin Xiang, Aiping Le, Jia-Ming Liu, Malik Peiris, Leo LM Poon, and Wei Zhang. Viral dynamics in mild and severe cases of covid-19. *The Lancet Infectious Diseases*, 2020.

[3] Ruiyun Li, Sen Pei, Bin Chen, Yimeng Song, Tao Zhang, Wan Yang, and Jeffrey Shaman. Substantial undocumented infection facilitates the rapid dissemination of novel coronavirus (sars-cov-2). *Science*, 368(6490):489–493, 2020.

[4] Derek K Chu, Elie A Akl, Stephanie Duda, Karla Solo, Sally Yaacoub, Holger J Schünemann, Amena El-harakeh, Antonio Bognanni, Tamara Lotfi, Mark Loeb, et al. Physical distancing, face masks, and eye protection to prevent person-to-person transmission of sars-cov-2 and covid-19: a systematic review and meta-analysis. *The Lancet*, 2020.

[5] Benjamin Rader, Laura F White, Michael R Burns, Jack Chen, Joseph Brilliant, Jon Cohen, Jeffrey Shaman, Larry Brilliant, Moritz UG Kraemer, Jared B Hawkins, et al. Mask-wearing and control of sars-cov-2 transmission in the usa: a cross-sectional study. *The Lancet Digital Health*, 2021.

[6] Michael Klompas, Charles A Morris, Julia Sinclair, Madelyn Pearson, and Erica S Shenoy. Universal masking in hospitals in the covid-19 era. *New England Journal of Medicine*, 382(21):e63, 2020.

[7] Trisha Greenhalgh, Manuel B Schmid, Thomas Czypionka, Dirk Bassler, and Laurence Gruer. Face masks for the public during the covid-19 crisis. *Bmj*, 369, 2020.

[8] Juliette Stehlé, Nicolas Voirin, Alain Barrat, Ciro Cattuto, Vittoria Colizza, Lorenzo Isella, Corinne Régis, Jean-François Pinton, Nagham Khanafer, Wouter Van den Broeck, et al. Simulation of an seir infectious disease model on the dynamic contact network of conference attendees. *BMC medicine*, 9(1):87, 2011.

[9] Julie Fournet and Alain Barrat. Epidemic risk from friendship network data: an equivalence with a non-uniform sampling of contact networks. *Scientific reports*, 6(1):1–11, 2016.

[10] Alain Barrat, Marc Barthelemy, and Alessandro Vespignani. *Dynamical processes on complex networks*. Cambridge university press, 2008.

[16] Peter Ashcroft, Jana S Huisman, Sonja Lehtinen, Judith A Bouman, Christian L Althaus, Roland R Regoes, and Sebastian Bonhoeffer. Covid-19 infectivity profile correction. *arXiv preprint arXiv:2007.06602*, 2020.

[17] Qun Li, Xuhua Guan, Peng Wu, Xiaoye Wang, Lei Zhou, Yeqing Tong, Ruiqi Ren, Kathy S.M. Leung, Eric H.Y. Lau, Jessica Y. Wong, Xuesen Xing, Nijuan Xiang, Yang Wu, Chao Li, Qi Chen, Dan Li, Tian Liu, Jing Zhao, Man Liu, Wenxiao Tu, Chuding Chen, Lianmei Jin, Rui Yang, Qi Wang, Suhua Zhou, Rui Wang, Hui Liu, Yinbo Luo, Yuan Liu, Ge Shao, Huan Li, Zhongfa Tao, Yang Yang, Zhiqiang Deng, Boxi Liu, Zhitao Ma, Yanping Zhang, Guoqing Shi, Tommy T.Y. Lam, Joseph T. Wu, George F. Gao, Benjamin J. Cowling, Bo Yang, Gabriel M. Leung, and Zijian Feng. Early transmission dynamics in wuhan, china, of novel coronavirus–infected pneumonia. *New England Journal of Medicine*, 382(13):1199–1207, 2020. PMID: 31995857.

[18] Piotr Sapiezynski, Arkadiusz Stopczynski, David Dreyer Lassen, and Sune Lehmann. Interaction data from the Copenhagen Networks Study. *Scientific Data*, 6(315), 2019.

[19] Christophe Fraser, Steven Riley, Roy M. Anderson, and Neil M. Ferguson. Factors that make an infectious disease outbreak controllable. *Proceedings of the National Academy of Sciences*, 101(16):6146–6151, 2004.

[20] Gabriel Kaptchuk, Eszter Hargittai, and Elissa M Redmiles. How good is good enough for covid19 apps? the influence of benefits, accuracy, and privacy on willingness to adopt. *arXiv preprint arXiv:2005.04343*, 2020.

[21] Jay J Van Bavel, Katherine Baicker, Paulo S Boggio, Valerio Capraro, Aleksandra Cichocka, Mina Cikara, Molly J Crockett, Alia J Crum, Karen M Douglas, James N Druckman, et al. Using social and behavioural science to support covid-19 pandemic response. *Nature Human Behaviour*, pages 1–12, 2020.

[22] Robert A Blair, Benjamin S Morse, and Lily L Tsai. Public health and public trust: Survey evidence from the ebola virus disease epidemic in liberia. *Social Science & Medicine*, 172:89–97, 2017.

[23] Patrick Vinck, Phuong N Pham, Kenedy K Bindu, Juliet Bedford, and Eric J Nilles. Institutional trust and misinformation in the response to the 2018–19 ebola outbreak in north kivu, dr congo: a population-based survey. *The Lancet Infectious Diseases*, 19(5):529–536, 2019.

[24] Anatol Rapoport, Albert M Chammah, and Carol J Orwant. *Prisoner's dilemma: A study in conflict and cooperation*, volume 165. University of Michigan press, 1965.

[36] Mirjam Kretzschmar, Ganna Rozhnova, and Michiel van Boven. Isolation and contact tracing can tip the scale to containment of covid-19 in populations with social distancing. *Available at SSRN 3562458*, 2020.

[37] Hossein Gorji, Markus Arnoldini, David F Jenny, Wolf-Dietrich Hardt, and Patrick Jenny. Stecc: Smart testing with contact counting enhances covid-19 mitigation by bluetooth app based contact tracing. *medRxiv*, 2020.

[38] Christophe Fraser, Lucie Abeler-Dörner, Luca Ferretti, Michael Parker, Michelle Kendall, and David Bonsall. Digital contact tracing: comparing the capabilities of centralised and decentralised data architectures to effectively suppress the COVID-19 epidemic whilst maximising freedom of movement and maintaining privacy. `https://github.com/BDI-pathogens/covid-19_instant_tracing`, 2020.

[39] Sadamori Kojaku, Laurent Hébert-Dufresne, and Yong-Yeol Ahn. The effectiveness of contact tracing in heterogeneous networks. *arXiv preprint arXiv:2005.02362*, 2020.

[40] Josh A Firth, Joel Hellewell, Petra Klepac, Stephen Kissler, Adam J Kucharski, and Lewis G Spurgin. Using a real-world network to model localized covid-19 control strategies. *Nature medicine*, pages 1–7, 2020.

[41] Lars Lorch, William Trouleau, Stratis Tsirtsis, Aron Szanto, Bernhard Schölkopf, and Manuel Gomez-Rodriguez. A spatiotemporal epidemic model to quantify the effects of contact tracing. *Testing, and Containment*, 2020.

[42] Alain Barrat, Ciro Cattuto, Mikko Kivelä, Sune Lehmann, and Jari Saramäki. Effect of manual and digital contact tracing on covid-19 outbreaks: a study on empirical contact data. *medRxiv*, 2020.

[43] Adam J Kucharski, Petra Klepac, Andrew JK Conlan, Stephen M Kissler, Maria L Tang, Hannah Fry, Julia R Gog, W John Edmunds, Jon C Emery, Graham Medley, et al. Effectiveness of isolation, testing, contact tracing, and physical distancing on reducing transmission of sars-cov-2 in different settings: a mathematical modelling study. *The Lancet Infectious Diseases*, 20(10):1151–1160, 2020.

[44] Jesús A Moreno López, Beatriz Arregui-García, Piotr Bentkowski, Livio Bioglio, Francesco Pinotti, Pierre-Yves Boëlle, Alain Barrat, Vittoria Colizza, and Chiara Poletto. Anatomy of digital contact tracing: role of age, transmission setting, adoption and case detection. *medRxiv*, 2020.

[45] Robert Hinch, Will Probert, Anel Nurtay, Michelle Kendall, Chris Wymant, Matthew Hall, Katrina Lythgoe, Ana Bulas Cruz, Lele Zhao, Andrea Stewart, Michael Ferretti, Luca Parker, Ares Meroueh, Bryn Mathias, Scott Stevenson, Daniel Montero, James Warren, Nicole K Mather, Anthony Finkelstein, Lucie Abeler-Dörner, and Christophe Bonsall, David Fraser. Effective configurations of a digital contact tracing app: A report to NHSX, 2020. `https://github.com/BDI-pathogens/covid-19_instant_tracing`.

[46] Matthew Abueg, Robert Hinch, Neo Wu, Luyang Liu, William JM Probert, Austin Wu, Paul Eastham, Yusef Shafi, Matt Rosencrantz, Michael Dikovsky, et al. Modeling the combined effect of digital exposure notification and non-pharmaceutical interventions on the covid-19 epidemic in washington state. *medRxiv*, 2020.

[47] Ying Mao, Susiyan Jiang, Daniel Nametz, Yuxin Lin, Jake Hack, John Hensley, Ryan Monaghan, and Tess Gutenbrunner. Data-driven analytical models of covid-2019 for epidemic prediction, clinical diagnosis, policy effectiveness and contact tracing: A survey, 2020.
